# The Cardiovascular Impact and Genetics of Pericardial Adiposity

**DOI:** 10.1101/2023.07.16.23292729

**Authors:** Joel T Rämö, Shinwan Kany, Cody R Hou, Samuel F Friedman, Carolina Roselli, Victor Nauffal, Satoshi Koyama, Juha Karjalainen, FinnGen, Mahnaz Maddah, Aarno Palotie, Patrick T Ellinor, James P Pirruccello

**Affiliations:** Cardiovascular Disease Initiative, Broad Institute of MIT and Harvard, Cambridge, MA, USA; Cardiovascular Research Center, Massachusetts General Hospital, Boston, MA, USA; Institute for Molecular Medicine Finland (FIMM), Helsinki Institute of Life Science (HiLIFE), University of Helsinki, Helsinki, Finland; Department of Cardiology, University Heart and Vascular Center Hamburg-Eppendorf, Hamburg, Germany; University of Minnesota Medical School, Minneapolis, Minnesota, USA; Broad Institute of MIT and Harvard, Cambridge, MA, USA; Department of Cardiology, University of Groningen, University Medical Center Groningen, Groningen, The Netherlands; Division of Cardiovascular Medicine, Brigham and Women’s Hospital, Boston, MA, USA; Analytic and Translational Genetics Unit, Massachusetts General Hospital and Harvard Medical School, Boston, MA, USA; Stanley Center for Psychiatric Research, Broad Institute of MIT and Harvard, Cambridge, MA, USA; Psychiatric and Neurodevelopmental Genetics Unit, Massachusetts General Hospital and Harvard Medical School, Boston, MA, USA; Department of Neurology, Massachusetts General Hospital, Boston, MA, USA; Cardiovascular Research Center, Massachusetts General Hospital, Harvard Medical School, Boston, MA, USA; Cardiology Division, Massachusetts General Hospital, Boston, MA, USA; Harvard Medical School, Boston, MA, USA; Bakar Computation Health Sciences Institute, University of California San Francisco, San Francisco, CA, USA; Division of Cardiology, University of California San Francisco, San Francisco, California, USA, San Francisco, CA, USA; Institute for Human Genetics, University of California San Francisco, San Francisco, CA, USA

## Abstract

**Background:** While previous studies have reported associations of pericardial adipose tissue (PAT) with cardiovascular diseases such as atrial fibrillation and coronary artery disease, they have been limited in sample size or drawn from selected populations. Additionally, the genetic determinants of PAT remain largely unknown. We aimed to evaluate the association of PAT with prevalent and incident cardiovascular disease and to elucidate the genetic basis of PAT in a large population cohort.

**Methods:** A deep learning model was trained to quantify PAT area from four-chamber magnetic resonance images in the UK Biobank using semantic segmentation. Cross-sectional and prospective cardiovascular disease associations were evaluated, controlling for sex and age. A genome-wide association study was performed, and a polygenic score (PGS) for PAT was examined in 453,733 independent FinnGen study participants.

**Results:** A total of 44,725 UK Biobank participants (51.7% female, mean [SD] age 64.1 [7.7] years) were included. PAT was positively associated with male sex (β = +0.76 SD in PAT), age (*r* = 0.15), body mass index (BMI; *r* = 0.47) and waist-to-hip ratio (*r* = 0.55) (P < 1×10^-230^). PAT was more elevated in prevalent heart failure (β = +0.46 SD units) and type 2 diabetes (β = +0.56) than in coronary artery disease (β = +0.22) or AF (β = +0.18). PAT was associated with incident heart failure (HR = 1.29 per +1 SD in PAT [95% CI 1.17–1.43]) and type 2 diabetes (HR = 1.63 [1.51–1.76]) during a mean 3.2 (±1.5) years of follow-up; the associations remained significant when controlling for BMI. We identified 5 novel genetic loci for PAT and implicated transcriptional regulators of adipocyte morphology and brown adipogenesis (*EBF1*, *EBF2* and *CEBPA*) and regulators of visceral adiposity (*WARS2* and *TRIB2*). The PAT PGS was associated with T2D, heart failure, coronary artery disease and atrial fibrillation in FinnGen (ORs 1.03–1.06 per +1 SD in PGS, P < 2×10^-10^).

**Conclusions:** PAT shares genetic determinants with abdominal adiposity and is an independent predictor of incident type 2 diabetes and heart failure.

**Clinical Perspective:** What is new?

- In a large, prospective and uniformly phenotyped cohort, pericardial adipose tissue was independently predictive of incident heart failure and type 2 diabetes when adjusted for body mass index.
- In contrast, pericardial adipose tissue was not independently predictive of atrial fibrillation.
- A genome-wide association study of pericardial adipose tissue identified five novel loci, implicating genes influencing adipocyte morphology, brown-like adipose tissue differentiation and abdominal adiposity.

What are the clinical implications?

- Pericardial adipose tissue accumulation may reflect a metabolically unhealthy adiposity phenotype similarly to abdominal visceral adiposity.

## Introduction

The increased global burden of obesity as a leading cause and modifiable risk factor for cardiovascular diseases is well recognized^1^. Not all obesity is alike, however, and the distribution of adipose tissue may be as important as its quantity. Abdominal adiposity, in particular, is associated with higher cardiovascular disease risk than subcutaneous adiposity^1,2^. Similarly, ectopic fat storage surrounding the heart has been suggested to confer independent cardiovascular risk^1^.

Pericardial adipose tissue (PAT) comprises epicardial adipose tissue (EAT)—located between the visceral pericardium and myocardium—and extrapericardial adipose tissue.^3–5^ In addition to the ectopic location of PAT within the thoracic cavity, the vascular supply of EAT derives from branches of the coronary arteries, similarly to the myocardium which is in immediate proximity to EAT with no separating fascia.^6^ EAT displays features of brown-like or beige adipose tissue and has been hypothesized to have a cardioprotective role via thermogeneration and supply of free fatty acids.^7^ However, multiple studies have suggested that EAT can also promote disease via secretion of pro-inflammatory and pro-fibrotic mediators^8^.

To date, more than 30 studies have reported the relation between PAT or EAT and a range of cardiovascular outcomes^3,4,9–11^. Meta-analyses have also demonstrated associations of EAT with myocardial infarction, coronary revascularization, atrial fibrillation, and cardiac death^11^. However, these studies have been limited in sample size and clinically heterogeneous as they have often been drawn from selected populations such as patients undergoing surgery or treatment for acute coronary syndromes.

The UK Biobank (UKB) is a large and deeply phenotyped population cohort with an ongoing cardiac magnetic resonance (CMR) imaging substudy.^12,13^ Coupled with advances in deep learning based annotation methods, this dataset enables the assessment of cardiovascular traits such as PAT at scale. Genotyping of UKB participants allows for the simultaneous evaluation of the heritable determinants of PAT in a sample that is many times larger than previous cohorts.

In this study, we quantify PAT in 44,725 UKB participants by using a deep learning model. We first assess the cross-sectional and prospective disease associations of PAT with and without adjustment for BMI. We then evaluate the genetic determinants of PAT in UKB and the independent FinnGen study.

## Methods

### Study design

We included participants from UKB and the FinnGen study. Primary analyses examining pericardial adipose tissue were conducted in UKB, and secondary analyses examining a polygenic score for PAT were conducted in FinnGen (**Supplementary Figure 1**).

UKB is a deeply phenotyped prospective population-level cohort which recruited approximately 500,000 participants aged 40–69 in the UK between 2006–2010^12,13^. A subset of participants within an imaging substudy underwent CMR with 1.5 T scanners (Magnetom Aera, Siemens Healthcare). This study has been conducted using the UKB Application Numbers #7089 and #17488 and was approved by the Mass General Brigham institutional review board (protocol 2003P001563).

FinnGen is a collection of prospective Finnish epidemiological and disease-based cohorts and hospital biobank samples linked to electronic health records (https://www.finngen.fi/en).^14^ A total of 453,733 participants from the FinnGen Data Freeze 11 were included in this study.

### Semantic segmentation of pericardial adipose tissue

Four-chamber images at random parts of the cardiac cycle from 250 randomly selected UKB CMR substudy participants were manually annotated by a physician (JTR). Segmentation maps were traced for pericardial adipose tissue and adjacent mediastinal structures including the cardiac chambers (**Supplementary Methods**). A total of 160 images were used for training, 40 images for validation and 50 images kept in a hold-out test set. A UNet based deep learning model from the fastai library v2.7.11 was constructed in PyTorch v1.13.1 using a ResNet50 encoder^15,16^. The fine-tuned model was used to infer segmentation of pericardial adipose tissue in the test set and all remaining UKB participants.^15,16^ Training parameters are detailed in the **Supplementary Methods**.

### Epidemiologic analyses

In UKB, the following cardiovascular diseases were defined using a combination of International Classification of Diseases (ICD) codes, self-report, and procedure codes (**Supplementary Table 2**): atrial fibrillation of flutter (AF), coronary artery disease (CAD), heart failure (HF), stroke and type 2 diabetes (T2D). The associations of prevalent cardiovascular diseases with PAT were tested using linear regression models including PAT as the outcome and age and sex as covariates. The associations of PAT with incident diseases were tested using Cox proportional hazards models with time from imaging to diagnosis or censoring as the outcome and PAT (standard deviation scaled or percentile-stratified), sex, and age as the predictors. BMI was included as an additional covariate in adjusted models. Participants with the corresponding disease at the time of imaging were excluded from incident disease analyses. Follow-up time was censored on September 30, 2021.

In FinnGen, cardiovascular diseases were defined using a combination of International Classification of Diseases (ICD) codes from specialist inpatient, outpatient and cause-of-death registries, procedure codes, and medication reimbursement codes (**Supplementary Methods**).

### Genotyping, imputation, and genetic quality control

We excluded UKB participants from genetic analyses if they had mismatch between reported and inferred sex, were outliers for heterozygosity or missingness, or had putative sex chromosome aneuploidy based on central quality control. Genotyped variants with MAF >1%, minor allele count >100, genotype missingness <5% and Hardy-Weinberg Equilibrium p-values >1×10^-15^ were included in regenie step 1, and further UKB analyses were performed for imputed variants with INFO > 0.3 and minor allele frequency >1%.

Sample QC, genotyping and quality control for FinnGen samples has been reported previously.^14^ The FinnGen genotype imputation protocol is available at: https://dx.doi.org/10.17504/protocols.io.xbgfijw.

### Genome-wide association studies

We performed a genome-wide association study for PAT using the additive genetic model implemented in regenie v3.2.5^17^. In addition to PAT, we performed new GWAS for height, weight, BMI, and WHR in the CMR substudy participants to ensure comparability for genomic correlation analyses.

### Heritability and genetic correlation analyses

Based on the summary statistics from the custom GWAS in the UKB imaging substudy, using LD Score Regression (LDSC) with HapMap3 variants and a European ancestry reference panel^18^, we calculated single nucleotide polymorphism heritability for PAT and genetic correlations between PAT and anthropometric measurements.

### Polygenic score analyses

We estimated PGS weights for PAT using the PRScs program in ‘auto’ mode based on a publicly available UKB European ancestry linkage disequilibrium panel and 1,117,404 HapMap3 variants^19^. PGSs were computed for all individuals in the FinnGen study. The associations of the PAT PGS with cardiovascular diseases were evaluated using logistic regression models with sex, age at the end of study follow-up or death, genomic principal components 1–5, and the genotyping array as basic covariates. BMI was included as an additional covariate in adjusted models.

### Additional computational and statistical software

Variant positions were lifted over from the GRCh37 build to the GRCh38 build for polygenic scoring in FinnGen using the UCSC liftOver tool^9^. All statistical analyses not otherwise specified were carried out in R (version 4.3.0)^10^.

## Results

### Semantic segmentation of PAT with deep learning

We began by manually annotating PAT in 200 four-chamber CMR images from randomly selected UKB participants at random phases of the cardiac cycle. We then fine-tuned a deep learning model based on ResNet50 to annotate PAT in the remaining participants. In a held out test set of 50 individuals, the model achieved a Dice score of 0.80 compared with a human annotator, similar to a recently reported model based on UKB CMR data.^5^ We further excluded 788 participants whose four-chamber images were not predicted to show at least 5 cm^2^ of each cardiac chamber as a quality control step to remove misaligned or poor-quality images (**Supplementary Methods**). The area of PAT (in cm^2^) was computed in all remaining 44,475 participants.

### The associations of PAT with demographic characteristics and anthropometric measures

In the study sample of 44,475 individuals, 51.7% of participants were female and the mean [SD] age was 64.1 [7.7] years (**Table 1**). The majority (96.7%) of the participants were of self-reported White ethnic background; participant characteristics by ethnic background are additionally reported in **Supplementary Table 1**. Men had on average more PAT compared with women (+0.78 SD units, P < 3e-324) (**Supplementary Figure 2**). Greater age was also associated with increased PAT (*r* = 0.15, P = 9.3×10^-229^).

**Table 1.**
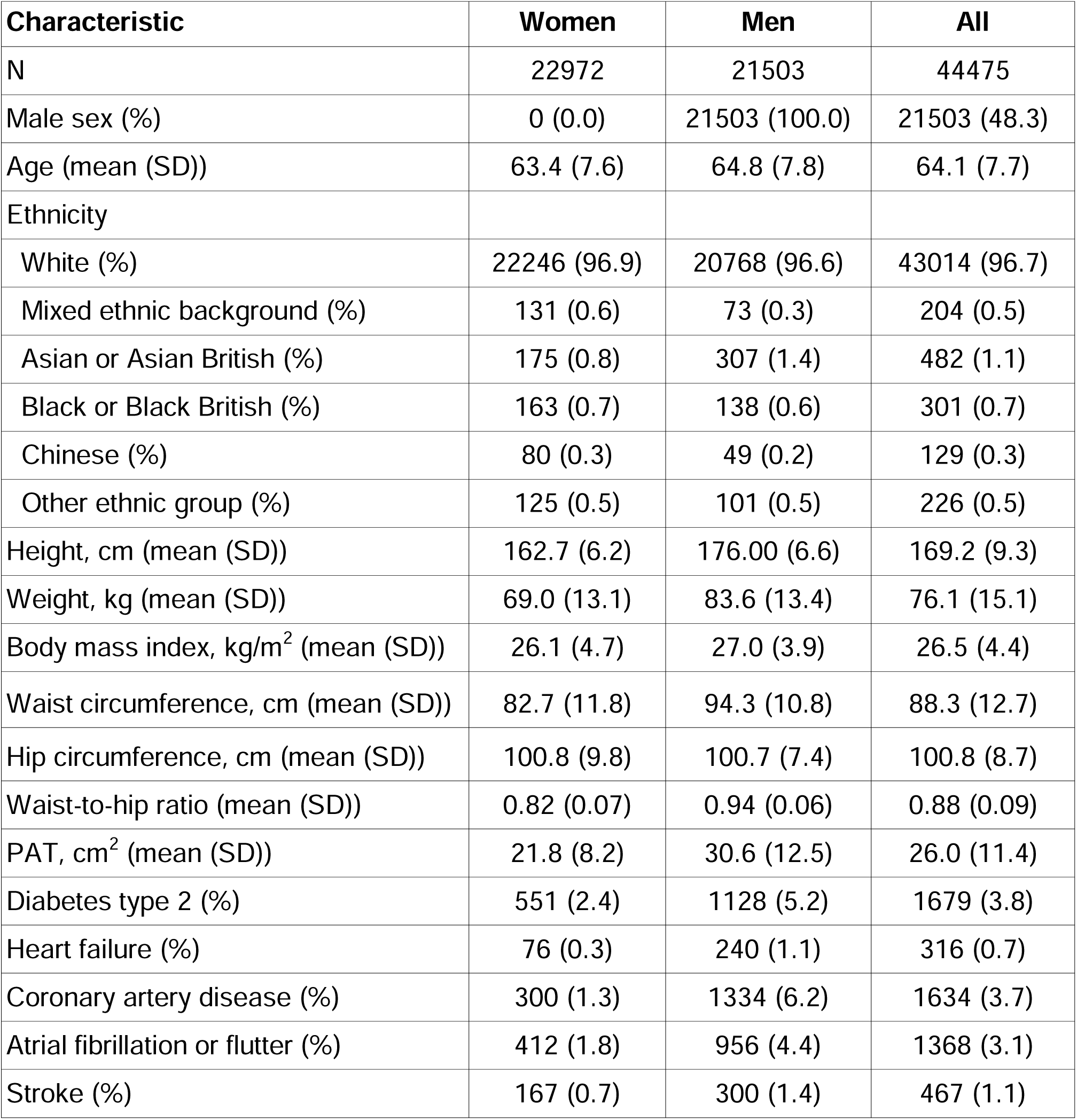
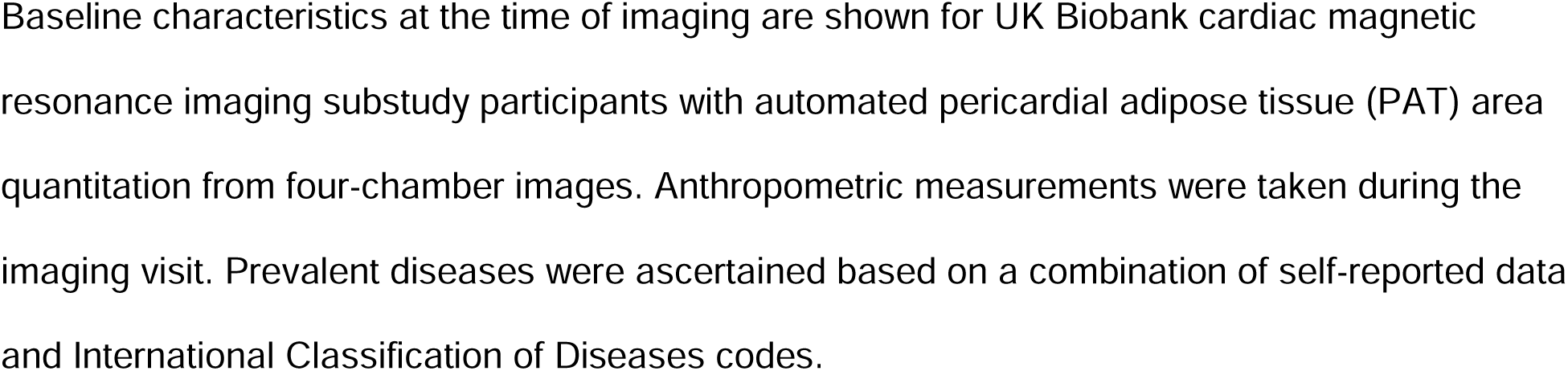
Characteristics of the study sample at the time of imaging.

**Table 2.** Lead variants from the genome-wide association study of pericardial adipose tissue in UK Biobank.

| Chr | Position | rsID | NEA | EA | Nearest Gene | Nearest Gene Annotation | Top PoPS gene | EAF | Beta | SE | P |
| --- | --- | --- | --- | --- | --- | --- | --- | --- | --- | --- | --- |
| 1 | 119699426 | rs7530762 | A | G | WARS2-AS1 | intronic | WARS2 | 0.67 | -0.62 | 0.075 | 2.4E-16 |
| 2 | 12882822 | rs16350 | ATC | A | TRIB2 | UTR3 | TRIB2 | 0.47 | 0.52 | 0.072 | 6.1E-13 |
| 3 | 49799046 | rs577934120 | CA | C | IP6K1 | intronic | RBM6 | 0.45 | 0.42 | 0.075 | 1.8E-08 |
| 5 | 55794632 | rs30351 | G | A | C5orf67 | intergenic | MIER3 | 0.74 | -0.46 | 0.081 | 2.0E-08 |
| 5 | 158009651 | rs553600342 | A | AG | EBF1 | intergenic | EBF1 | 0.24 | -0.60 | 0.086 | 2.3E-12 |
| 8 | 25464670 | rs73221948 | G | T | CDCA2 | intergenic | EBF2 | 0.29 | -0.86 | 0.082 | 6.7E-26 |
| 19 | 34015904 | rs41119 | G | T | PEPD | intergenic | CEBPA | 0.36 | -0.42 | 0.076 | 3.7E-08 |

Variant positions are in the GRCh37 genome build. The nearest gene and the gene most strongly prioritized by the Polygenic Priority Score (PoPS) are shown for all lead variants. AF = allele frequency, Chr = chromosome, EA = effect allele, EAF = effect allele frequency, NEA = non-effect allele, PoPS = Polygenic Priority Score, SE = standard error.

We observed modest correlations with traditional anthropometric measures suggesting that PAT may convey additional information: PAT was associated with height (*r* = 0.31), weight (*r* = 0.57), BMI (*r* = 0.47) and WHR (*r* = 0.55) (P < 3×10^-324^ for all). Simple linear estimates based on WHR tended to underestimate PAT particularly in participants with high WHR and PAT.

### Prevalent cardiovascular diseases and PAT

We evaluated the associations of PAT with five prevalent cardiovascular diseases at the time of imaging (**Figure 2** and **Supplementary Table 3**). PAT was significantly elevated in individuals with prevalent T2D (+0.56 SD units in PAT, P = 6.9×10^-133^), HF (+0.46 SD, P = 7.0×10^-19^), CAD (+0.22 SD, P = 1.3×10^21^), and AF (+0.18 SD, P = 1.2×10^-2^), but not in individuals with prevalent stroke (+0.09 SD, P = 0.08).

**Figure 1.**
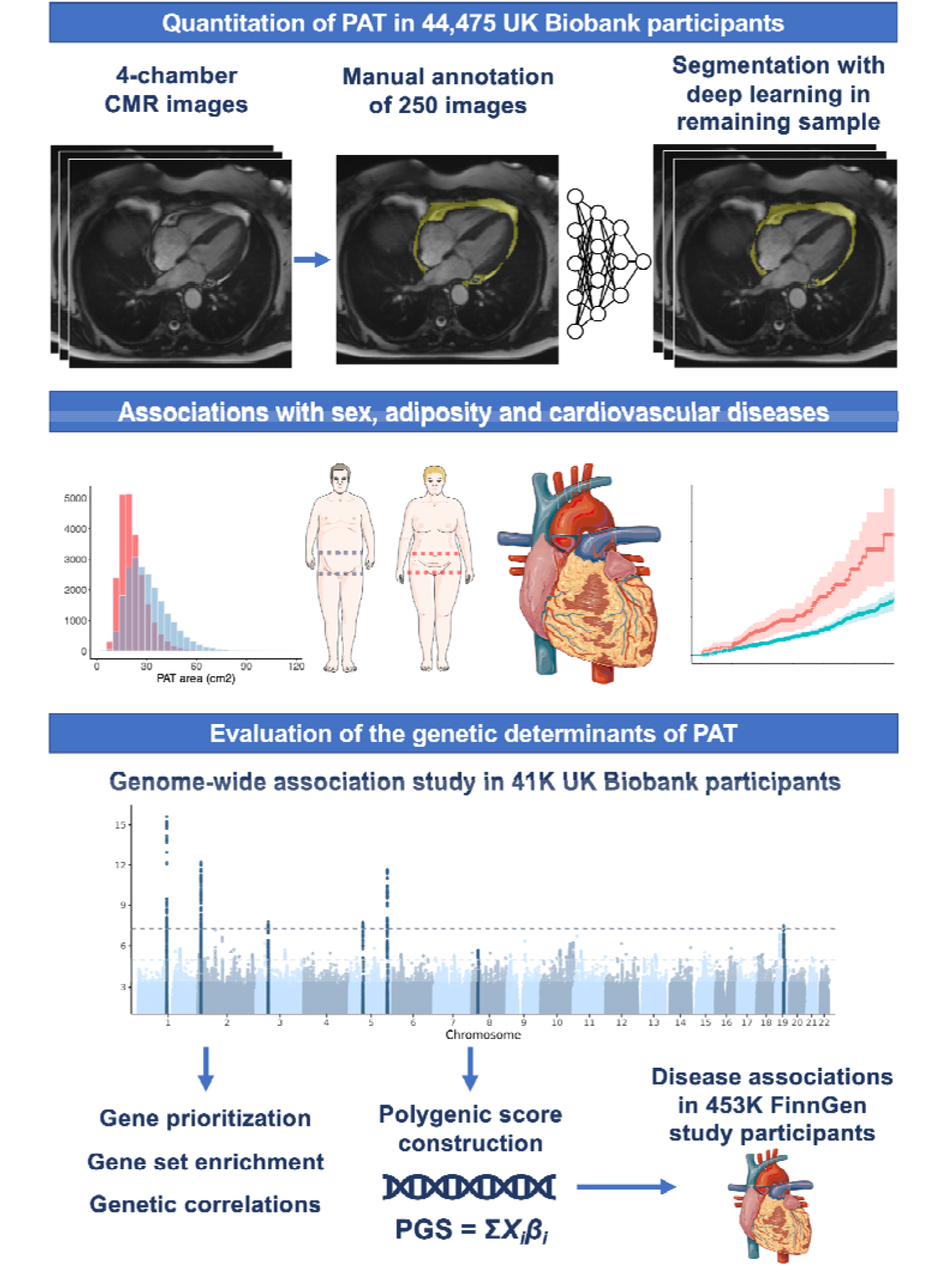
Automated quantitation of pericardial adipose tissue in 44,475 UK Biobank participants. Pericardial adipose tissue (PAT) was quantified with deep learning in 44,475 UK Biobank participants based on a separate training and test set of 250 4-chamber cardiac magnetic resonance images. The associations between PAT and baseline characteristics (at the time of imaging) and incident diseases were evaluated. A genome-wide association study of PAT was conducted in 41,494 participants and the resulting summary statistics were used in gene prioritization, gene set enrichment and genetic correlation analyses. The disease associations of a polygenic score for PAT were evaluated in 453,733 participants of the FinnGen study. Magnetic resonance images are reproduced by kind permission of UK Biobank ©. Parts of the figures were generated using Servier Medical Art, provided by Servier, licensed under a Creative Commons Attribution 3.0 unported license.

**Figure 2:**
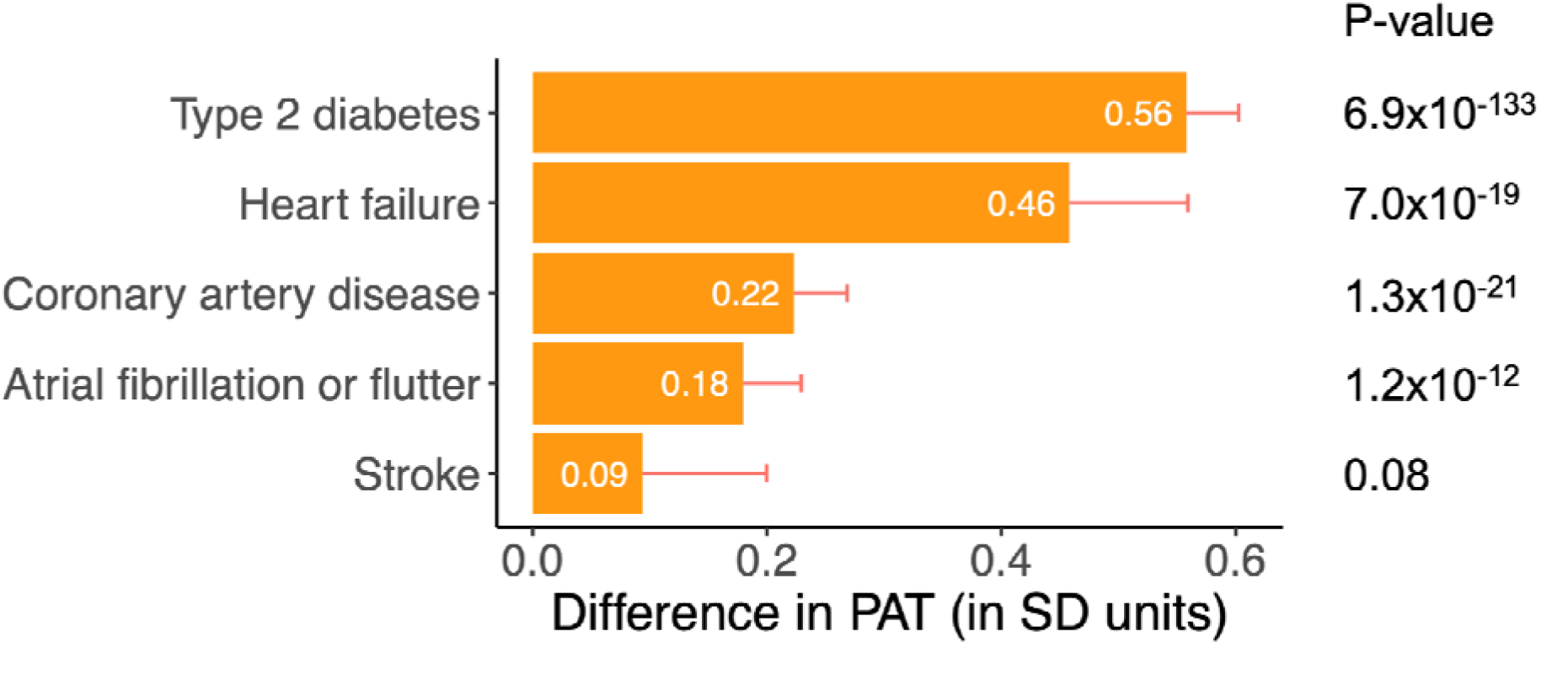
The associations of prevalent cardiovascular diseases with pericardial adipose tissue. The associations of prevalent diseases with pericardial adipose tissue (PAT) were examined with linear regression models including the respective disease, age, and sex as predictors and PAT as the outcome. Each bar corresponds to the difference in PAT (in SD units) between those with prevalent disease and those without. Error bars correspond to positive 95% confidence intervals.

### Predictive utility of PAT for incident cardiovascular diseases

In survival analyses, after excluding participants with prevalent diseases at the time of imaging, PAT as a continuous measurement was associated with incident T2D (HR 1.63 per +1 SD increment in PAT, 95% CI 1.51-1.76, P = 7.2×10^-36^), HF (HR 1.29, 95% CI 1.17–1.43, P = 4.8×10^-7^) and AF (HR 1.17, 95% CI 1.08–1.26, P = 4.6×10^-5^) (**Supplementary Figure 3** and **Supplementary Table 4**). We did not observe significant associations between PAT and incident coronary artery disease (HR 1.10, 95% CI 1.00-1.20, P = 0.052) or stroke (HR 1.12, 95% CI 0.99-1.27, P = 0.063). When including BMI as an additional covariate, PAT remained an independent predictor of incident T2D (HR 1.25, 95% CI 1.14–1.38, P = 1.6e-6) and HF (HR 1.16, 95% CI 1.03–1.31, P = 0.013) (**Supplementary Figure 3**).

Recapitulating the patterns in analyses of PAT as a continuous measurement, in analyses stratified by PAT decile (**Figure 3**, **Supplementary Figures 4–5**, and **Supplementary Table 5**), those in the highest 10% vs. those in lowest 0-90% of PAT had significantly elevated risk of incident T2D (HR 3.14, 95% CI 2.50–3.95, P = 1.5×10^-22^), HF (HR 1.91, 95% CI 1.41–2.57, P = 2.4×10^-5^) and AF (HR 1.47, 95% CI 1.17–1.84, P = 8.1×10^-4^), but no significant elevation in risk of of CAD (HR 1.33, 95% CI 0.99–1.77, P = 0.057) or stroke (HR 1.22, 95% CI 0.83-1.78, P = 0.31). Only the associations with incident T2D and HF remained significant when adjusted for BMI.

**Figure 3:**
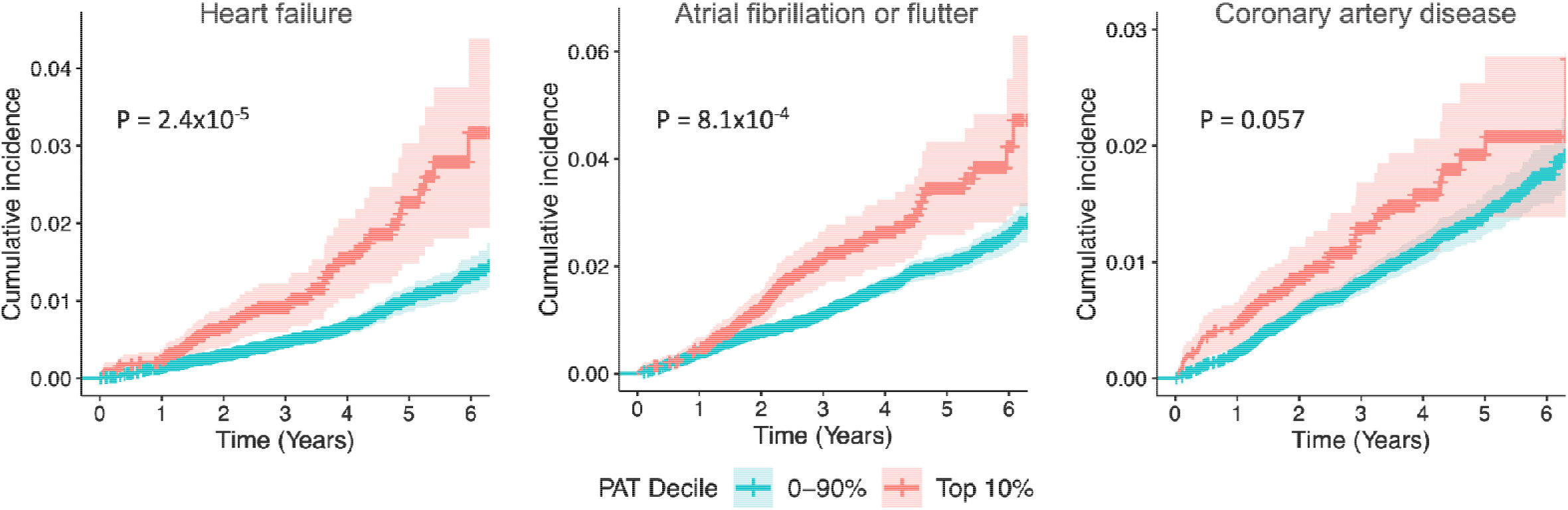
The associations of pericardial adipose tissue with incident cardiovascular diseases. Cumulative incidence curves for cardiovascular diseases are shown for participants stratified by PAT deciles at the time of imaging (top 10% vs others). The shaded areas correspond to 95% confidence intervals. Individuals with the corresponding disease at the time of imaging were excluded from the incident disease analyses. P-values were estimated using Cox Proportional hazards models with age and sex as additional covariates.

### Genome-wide association study of PAT

We performed a genome-wide association study of PAT in 41,494 UKB participants who passed genotyping quality control, and identified seven loci at genome-wide significance (P < 5e-8) (**Figure 4**, **Supplementary Results** and **Supplementary Figure 7**). These included the two previously reported loci for PAT (with lead variants in or near *TRIB2* and *EBF1*) and five novel PAT loci with lead variants in or near *CDCA2*, *WARS2*, *IP6K1*, *C5orf67*/*ANKRD55*, and *PEPD* (**Supplementary Figure 6**). The single nucleotide polymorphism based heritability of PAT on the observed scale was 0.15 (SE 0.02).

**Figure 4:**
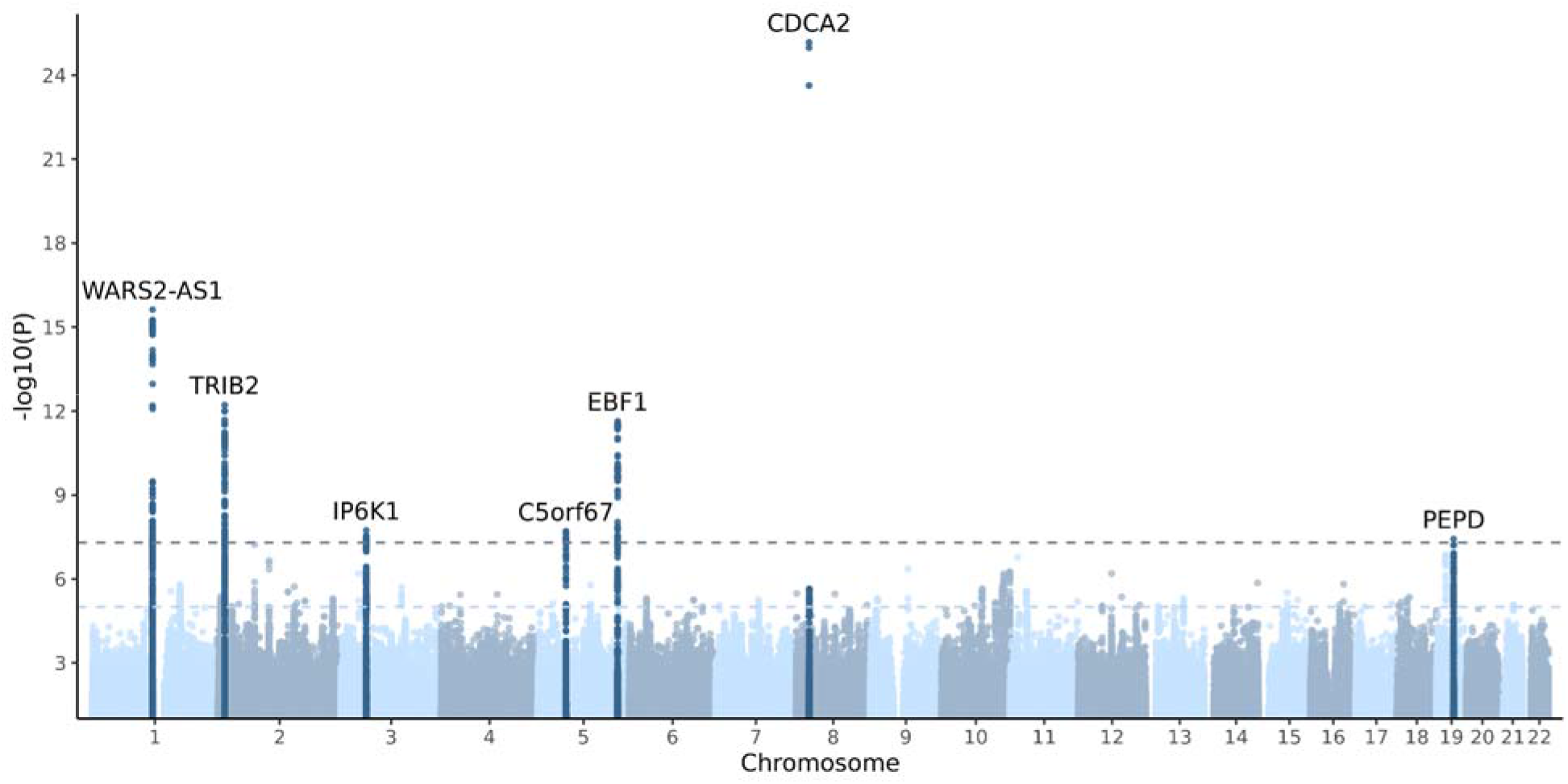
Manhattan plot of the genome-wide association study of pericardial adipose tissue in UK Biobank. A genome-wide association study of pericardial adipose tissue was performed in 41,494 UK Biobank participants. Each variant is plotted as a data point, with the corresponding -log_10_(P) shown on the y-axis and the genomic position shown on the x-axis grouped by chromosomes. The genome-wide significance threshold (P = 5×10^-8^) is shown with a darker dashed line and a suggestive threshold (P = 1×10^-5^) is shown with a lighter dashed line. Genomic loci with at least one variant reaching genome-wide significance are labeled with the name of the nearest gene, and all variants within 500 kilobases of the lead variant are colored in a darker blue for visualization purposes. The y-axis is truncated to only show variants with a P-value ≤ 0.1.

We used the nearest protein-coding gene and Polygenic Priority Score (PoPS) approaches to prioritize likely causal genes in the seven association loci^20^. The genes *WARS2*, *TRIB2*, *RBM6*, *MIER3*, *EBF1*, *EBF2*, and *CEBPA* were most strongly prioritized by PoPS in their respective loci (**Supplementary Table 6**). The nearest-gene approach and PoPS were concordant for *WARS2*, *TRIB2*, and *EBF1*.

Recapitulating correlations between measured PAT and many anthropometric variables, we also observed moderate genetic correlations between PAT and weight (*r*_g_ = 0.48, SE = 0.04, P = 5.9×10^-30^), BMI (*r*_g_ = 0.50, SE = 0.04, P = 4.3×10^-30^), and WHR (*r*_g_ = 0.60, SE = 0.05, P = 2.7×10^-30^) (**Supplementary Table 7**). In contrast, PAT was not genetically correlated with measured height (*r*_g_ = 0.05, P = 0.31).

### Meta-analysis of genome-wide association studies of PAT

We additionally performed a meta-analysis of the UKB GWAS and a previously published smaller GWAS meta-analysis (n = up to 12,204)^21^. This meta-analysis of up to 53,698 participants was conducted based on p-values and effect direction due to the lack of effect size estimates in the previous meta-analysis that incorporated PAT indices from different imaging modalities (**Supplemental Methods**). In the new meta-analysis, we identified one novel locus (*CYP26B1*). The *CDCA2* locus did not reach significance, potentially because the three closely clustering variants driving the association in UKB were not included in the previously published meta-analysis and could not be analyzed together. (**Supplementary Figures 8–9** and **Supplementary Table 8**).

### Predictive utility of a polygenic score for PAT

Lastly, we constructed a genome-wide polygenic score (PGS) for PAT using the PRS-CS method based on the summary statistics from the UKB GWAS. We subsequently evaluated the disease associations of the PAT PGS in 453,733 participants of the independent FinnGen study, sample characteristics of which are shown in **Supplementary Table 9**^14^. In logistic regressions, the PAT PGS was significantly associated with T2D, HF, CAD, AF or flutter, and stroke (P < 0.004 for all) (**Figure 5A** and **Supplementary Table 10**). These positive associations remained significant even when including BMI as an additional covariate (P < 0.007 for all) (**Figure 5B** and **Supplementary Table 10**).

**Figure 5:**
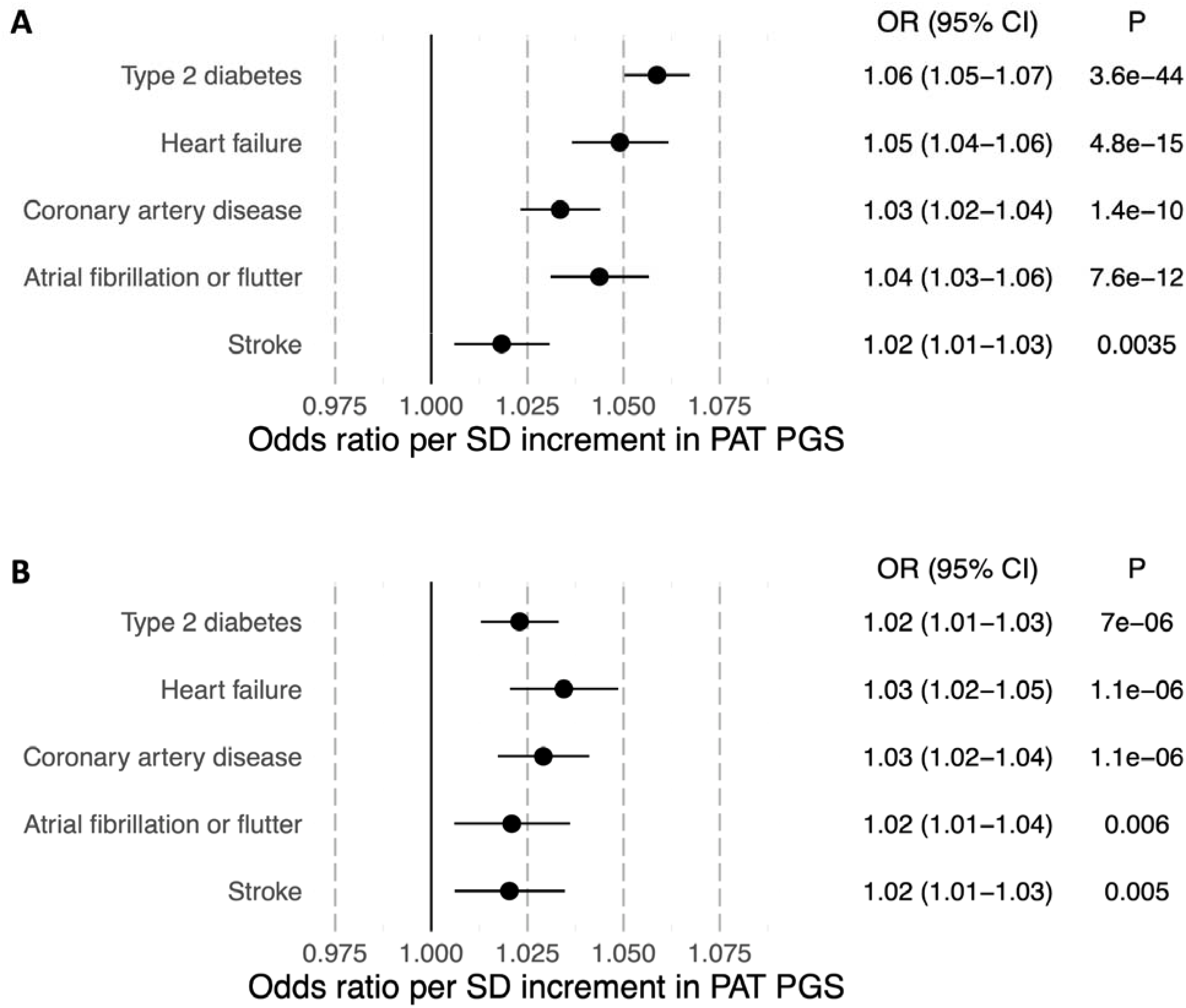
The predictive utility of a polygenic score for pericardial adipose tissue in FinnGen. A polygenic score (PGS) for pericardial adipose tissue (PAT) was constructed using PRS-CS based on summary statistics from the genome-wide association study of PAT in UK Biobank and subsequently applied to 453,733 participants in the FinnGen study. A. The associations of the PAT PGS with cardiovascular diseases were evaluated using logistic regression models with sex, age at the end of study follow-up or death, genomic principal components 1–5 and the genotyping array as basic covariates. B. The associations of the PAT PGS with cardiovascular diseases were examined including body mass index (BMI) as an additional covariate. Odds ratios are shown per SD increment in PAT PGS.

## Discussion

In this study, we examined the cardiovascular associations and genetic determinants of PAT in more than 40,000 individuals from a large, prospective and uniformly phenotyped cohort. These analyses enable several insights. We expand prior knowledge linking PAT as an independent predictor to HF and T2D. Genetic loci suggest that variation in PAT is influenced by regulators of adipocyte morphology, brown-like adipose tissue differentiation and abdominal adiposity. Our findings are consistent with PAT as a thoracic fat depot reflective of metabolically unhealthy adiposity independent of BMI.

Considerable interest has been focused on EAT as a potential local driver of cardiovascular disease. EAT has been hypothesized to contribute to AF by local secretion of profibrotic or inflammatory factors^8^. Paracrine or vasocrine release of cytokines and immune response factors from EAT to coronary diseases has been suggested to drive the development of atherosclerosis^7^. PAT—a superset that includes extrapericardial adipose in addition to EAT— has also been associated with visceral adipose tissue, as well as similar profiles of cardiovascular risk factors and metabolic syndrome^5,22^, which raises the question of whether many of the observed disease associations may reflect global consequences of unhealthy visceral adiposity rather than more localized cardiac effects.

Here, we identify PAT to be most predictive of incident T2D and HF even after controlling for BMI. Our findings are in keeping with recently reported associations with incident T2D in 42,598 UKB participants and with incident heart failure with preserved ejection fraction in 6,785 participants of the Multi-Ethnic Study of Atherosclerosis (MESA)^4^. We observed a positive although non-significant association with incident CAD, which was attenuated when controlling for BMI. Significant independent predictive utility of PAT for incident CAD has previously been reported in a study of 6,814 MESA participants^3^. Finally, while we replicate an association between PAT and prevalent AF^9^, PAT had no predictive utility for AF when controlling for BMI, in keeping with an analysis of 7,991 participants from MESA and Jackson Heart Study^23^. In combination, these patterns of prospective associations are consistent with metabolically unhealthy adiposity.

We replicated two known genetic loci and identified five novel loci for PAT. Previously, a meta-analysis of 11,596 participants from heterogeneous imaging cohorts reported genetic loci containing *TRIB2* and *EBF1*, but provided no effect estimates for further assessments^21^. Here, the expansion of association loci allows several biological insights. A prominent finding is considerable genetic correlation and locus overlap with abdominal adiposity. Among genomic loci linked with PAT in this study, all have been previously associated with WHR^24^. Four of seven loci (*IP6K1*, *CDCA2*, *C5orf67*, and *PEPD*) have been associated with visceral adipose tissue, while the *WARS2*, *TRIB2* and *EBF1* loci may be more specific to PAT^25^. We also demonstrated that a polygenic score for PAT, when tested in the independent FinnGen study, recapitulated the disease-specific patterns observed in prospective UKB analyses, lending further validity to the phenotype definitions and suggesting that shared genetic variation affects both PAT and cardiovascular morbidity.

Prioritization of potentially causal genes in the association loci highlights interconnected biological pathways influencing PAT accumulation. On separate chromosomes, we prioritize the transcription factor encoding genes *EBF1* and *EBF2*. *EBF1* is a regulator of adipose cell morphology and lipolysis^26^, and decreased levels of *EBF1* have been observed in white adipose tissue hypertrophy. *EBF2* is a promoter of brown-like / beige adipose cell differentiation^27^. *CEBPA*, prioritized in another locus, encodes for the transcription factor CCAAT/enhancer binding protein alpha (C/EBPα), which shares binding sites with Peroxisome proliferator-activated receptor gamma (PPARγ) and acts as a co-stimulator of adipogenesis and adipocyte differentiation^28,29^. In a previously identified PAT locus, we also prioritize *TRIB2*, which is a promoter of CCAAT/enhancer binding protein beta, which transactivates the expression of both C/EBPα and PPARγ^30^. Finally, *WARS2* encodes the mitochondrial trytophanyl-tRNA synthetase; a mutant *Wars2* mouse model exhibits reduced food intake, resistance to diet-induced obesity, and changes in relative visceral adiposity.^31^ Although many of the association loci overlap with more general measures of adiposity, they may represent a targeted subset of drivers of unhealthy adiposity, unlike many loci linked with BMI that may exert their effects via neuronal pathways and hunger regulation^32^.

The findings should be interpreted in the context of the study design. Firstly, in our quantitation of PAT we were not able to differentiate between epicardial and paracardial (extrapericardial) adipose tissue. Secondly, PAT quantitation was based on a single CMR slice. The relative localization of PAT surrounding the heart may carry added significance. Thirdly, the accuracy of PAT quantitation was not perfect. However, reduced accuracy is more likely to predispose to type II rather than type I error, and the identification of known and novel GWAS loci suggests that the increased sample size still outweighs reduced accuracy for genomic discovery. Fourth, participants in the UKB are healthier than the overall population^33^, which may affect the external validity of disease risk estimates. Lastly, UKB participants were mostly of European ancestry, which may limit the generalizability of the findings to other ancestries.

In conclusion, we identify PAT as an independent predictor of incident T2D and HF. Individual variation in PAT is likely influenced by genes regulating abdominal adiposity, adipocyte morphology and brown-like adipogenesis. The intrathoracic accumulation of PAT may reflect a metabolically unhealthy adiposity phenotype similar to abdominal visceral adiposity.

## Non-standard Abbreviations and Acronyms

EAT: epicardial adipose tissue
GWAS: genome-wide association study
PAT: pericardial adipose tissue
PoPS: polygenic priority score

## Appendices

## Data availability

UK Biobank data are made available to researchers from research institutions with genuine research inquiries, following IRB and UK Biobank approval. GWAS summary statistics and polygenic score weights will be made available upon publication at the Broad Institute Cardiovascular Disease Knowledge Portal (http://www.broadcvdi.org). Pericardial adipose tissue measurements will be returned to the UK Biobank for use by any approved researcher. The Finnish biobank data can be accessed through the Fingenious® services (https://site.fingenious.fi/en/) managed by FINBB. Finnish Health register data can be applied for from Findata (https://findata.fi/en/data/).

## Acknowledgements

We want to acknowledge the participants and investigators of UK Biobank and FinnGen studies. The following biobanks are acknowledged for delivering biobank samples to FinnGen: Auria Biobank (www.auria.fi/biopankki), THL Biobank (www.thl.fi/biobank), Helsinki Biobank (www.helsinginbiopankki.fi), Biobank Borealis of Northern Finland (https://www.ppshp.fi/Tutkimus-ja-opetus/Biopankki/Pages/Biobank-Borealis-briefly-in-English.aspx), Finnish Clinical Biobank Tampere (www.tays.fi/en-US/Research_and_development/Finnish_Clinical_Biobank_Tampere), Biobank of Eastern Finland (www.ita-suomenbiopankki.fi/en), Central Finland Biobank (www.ksshp.fi/fi-FI/Potilaalle/Biopankki), Finnish Red Cross Blood Service Biobank (www.veripalvelu.fi/verenluovutus/biopankkitoiminta), Terveystalo Biobank (www.terveystalo.com/fi/Yritystietoa/Terveystalo-Biopankki/Biopankki/) and Arctic Biobank (https://www.oulu.fi/en/university/faculties-and-units/faculty-medicine/northern-finland-birth-cohorts-and-arctic-biobank). All Finnish Biobanks are members of BBMRI.fi infrastructure (www.bbmri.fi). Finnish Biobank Cooperative -FINBB (https://finbb.fi/) is the coordinator of BBMRI-ERIC operations in Finland.

## Author contributions

JPP, PTE, AP, and JTR conceived of the study. JTR annotated images. JTR, JPP and CRH trained the deep learning models. JTR conducted bioinformatic analyses. CRH, SK, SFF, CR, VN, SK, JK, and MM, provided analysis support. JTR, JPP, PTE, and SK wrote the paper. All authors contributed to the analysis plan or provided critical revisions.

