## Supplementary Information for "The Cardiovascular Impact and Genetics of Pericardial Adiposity"

#### Table of Contents


**Table of Contents** 1
**Supplementary Methods** 3
Fine-tuning of the deep learning model 3
Additional quality control for CMR images 3
FinnGen Ethics Statement 3
Endpoint definitions in FinnGen 5
Gene prioritization 5
Genome-wide association study meta-analysis 6
Polygenic score analyses 6
**Supplementary Results** 7
Genomic inflation and LD Score Regression Intercept 7
**Supplementary Figures** 8
Supplementary Figure 1: Study flowchart 8
Supplementary Figure 2: The associations of pericardial adipose tissue with age, sex, and anthropometric measurements 9
Supplementary Figure 3: The associations of pericardial adipose tissue with incident diseases 10
Supplementary Figure 4: The associations of percentile-stratified pericardial adipose tissue with incident diseases 11
Supplementary Figure 5: Cumulative incidences of cardiovascular diseases stratified by PAT decile at baseline 13
Supplementary Figure 6: Locus plots from the genome-wide association study of pericardial adipose tissue in UK Biobank 18
Supplementary Figure 7: Quantile-quantile plot of p-values from the genome-wide association study of pericardial adipose tissue in UK Biobank 20
Supplementary Figure 8: Manhattan plot of the meta-analysis of genome-wide association studies of pericardial adipose tissue 21
Supplementary Figure 9: Quantile-quantile plot of the meta-analysis of genome-wide association studies of pericardial adipose tissue 22
**Supplementary Tables** 23
Supplementary Table 1: Characteristics of the study sample at the time of imaging and stratified by self-reported ethnic background 23
Supplementary Table 2: UK Biobank Disease Definitions 24

Supplementary Table 3: Prevalent disease association results in the UK Biobank 27
Supplementary Table 4: The associations of pericardial adipose tissue with incident diseases 28
Supplementary Table 5: The associations of percentile-stratified pericardial adipose tissue with incident diseases 29
Supplementary Table 6: Polygenic priority scores for genes in genome-wide significant loci in the genome-wide association study of pericardial adipose tissue in UK Biobank 30
Supplementary Table 7: Genetic correlations between pericardial adipose tissue and anthropometric traits 33
Supplementary Table 8: Lead variants from the meta-analysis of genome-wide association studies of pericardial adipose tissue 34
Supplementary Table 9: FinnGen participant characteristics 35
Supplementary Table 10: The predictive utility of a polygenic score for pericardial adipose tissue in FinnGen 36
**Supplementary References** **37**

### Supplementary Methods

#### Fine-tuning of the deep learning model

All four-chamber images were resized to 168 x 208 pixels and normalized using mean and standard deviation estimates from ImageNet[1]. The model was fine-tuned with a batch size of four. Augmentations with rotations up to 90 degrees were applied. Gradient clipping was used to limit the size of the gradients. The learning rate was chosen as $1x10^{-4}$ based on the learning rate finder and the model was trained for 50 cycles while monitoring the Dice score. We applied the fine-tuned model to randomly selected four-chamber CMR images from all remaining 45,263 UKB participants after excluding participants in the training, validation and test sets.

#### Additional quality control for CMR images

A small proportion of the UK Biobank four-chamber CMR images were misaligned (not true four-chamber images) or of poor quality e.g. due to imaging artifacts. To exclude these images from final analyses, we first manually annotated the four cardiac chambers (left ventricle, left atrium, right ventricle, right atrium) in the same training data that was used for PAT model training. The deep learning model based on Resnet50 was fine-tuned with additional labels for the four cardiac chambers and used to infer the segmentation of the cardiac chambers in addition to PAT[2]. We excluded any images in which any of the cardiac chambers was predicted to have an area less than $5cm^2$.

#### FinnGen Ethics Statement

Patients and control subjects in FinnGen provided informed consent for biobank research, based on the Finnish Biobank Act. Alternatively, separate research cohorts, collected prior the Finnish Biobank Act came into effect (in September 2013) and start of FinnGen (August 2017), were collected based on study-specific consents and later transferred to the Finnish biobanks after approval by Fimea (Finnish Medicines Agency), the National Supervisory Authority for Welfare and Health. Recruitment protocols followed the biobank protocols approved by Fimea. The Coordinating Ethics Committee of the Hospital District of Helsinki and Uusimaa (HUS) statement number for the FinnGen study is Nr HUS/990/2017.

The FinnGen study is approved by Finnish Institute for Health and Welfare (permit numbers: THL/2031/6.02.00/2017, THL/1101/5.05.00/2017, THL/341/6.02.00/2018, THL/2222/6.02.00/2018, THL/283/6.02.00/2019, THL/1721/5.05.00/2019 and THL/1524/5.05.00/2020), Digital and population data service agency (permit numbers: VRK43431/2017-3, VRK/6909/2018-3, VRK/4415/2019-3), the Social Insurance Institution (permit numbers: KELA 58/522/2017, KELA 131/522/2018, KELA 70/522/2019, KELA 98/522/2019, KELA 134/522/2019, KELA 138/522/2019, KELA 2/522/2020, KELA 16/522/2020), Findata permit numbers THL/2364/14.02/2020, THL/4055/14.06.00/2020, THL/3433/14.06.00/2020, THL/4432/14.06/2020, THL/5189/14.06/2020, THL/5894/14.06.00/2020, THL/6619/14.06.00/2020, THL/209/14.06.00/2021, THL/688/14.06.00/2021, THL/1284/14.06.00/2021, THL/1965/14.06.00/2021, THL/5546/14.02.00/2020, THL/2658/14.06.00/2021, THL/4235/14.06.00/2021, Statistics Finland (permit numbers: TK-53-1041-17 and TK/143/07.03.00/2020 (earlier TK-53-90-20) TK/1735/07.03.00/2021, TK/3112/07.03.00/2021) and Finnish Registry for Kidney Diseases permission/extract from the meeting minutes on 4$^{th}$ July 2019.

The Biobank Access Decisions for FinnGen samples and data utilized in FinnGen Data Freeze 10 include: THL Biobank BB2017_55, BB2017_111, BB2018_19, BB_2018_34, BB_2018_67, BB2018_71, BB2019_7, BB2019_8, BB2019_26, BB2020_1, BB2021_65, Finnish Red Cross Blood Service Biobank 7.12.2017, Helsinki Biobank HUS/359/2017, HUS/248/2020, HUS/150/2022 § 12, §13, §14, §15, §16, §17, §18, and §23, Auria Biobank AB17-5154 and amendment #1 (August 17 2020) and amendments BB_2021-0140, BB_2021-0156 (August 26 2021, Feb 2 2022), BB_2021-0169, BB_2021-0179, BB_2021-0161, AB20-5926 and amendment #1 (April 23 2020)and it´s modification (Sep 22 2021), Biobank Borealis of Northern Finland_2017_1013, 2021_5010, 2021_5018, 2021_5015, 2021_5023, 2021_5017, 2022_6001, Biobank of Eastern Finland 1186/2018 and amendment 22 § /2020, 53§/2021, 13§/2022, 14§/2022, 15§/2022, Finnish Clinical Biobank Tampere MH0004 and amendments (21.02.2020 & 06.10.2020), §8/2021, §9/2022, §10/2022, §12/2022, §20/2022, §21/2022, §22/2022, §23/2022, Central Finland Biobank 1-2017, and Terveystalo Biobank STB 2018001 and amendment 25$^{th}$ Aug 2020, Finnish Hematological Registry and Clinical Biobank decision 18$^{th}$ June 2021, Arctic biobank P0844: ARC_2021_1001.

#### Endpoint definitions in FinnGen

We used the following FinnGen endpoints from Data Freeze 11: E4_DM2 (Type 2 diabetes), I9_AF (atrial fibrillation or flutter), I9_HEARTFAIL (heart failure), I9_CHD (Coronary artery disease), and I9_STR (stroke). FinnGen endpoint definitions incorporate International Classification of Diseases (ICD) codes from hospital, outpatient and cause-of-death registries; ICD or Social Insurance Institution (KELA) classification based medication reimbursement codes; and Anatomical Therapeutic Chemical codes for medication purchases. The definitions are publicly available at https://www.finngen.fi/en/researchers/clinical-endpoints and on https://risteys.finregistry.fi.

#### Gene prioritization

We used two approaches to prioritize potentially causative genes in the association loci: nearest protein-coding gene and Polygenic Priority Score (PoPS)[3]. To compute gene-specific PoPS, we first performed a gene-based analysis with MAGMA v1.10 using the default snp-wise=mean gene analysis model[4]. The raw gene-based results from MAGMA and the full set of default features provided by the PoPS authors were then used to calculate gene-specific PoPS. The gene with the highest PoPS within +/- 500kb of the lead variant was designated as the gene prioritized by PoPS.

#### Genome-wide association study meta-analysis

We performed a p-value based sample size weighted meta-analysis of the UKB GWAS and a previously published GWAS meta-analysis (n = 12,204)[5,6] using METAL (version release 2020-05-05) without genomic control. We evaluated for the presence of inflation based on the LD score regression intercept for the novel meta-analysis.

#### Polygenic score analyses

We constructed a polygenic score (PGS) for PAT using the summary statistics from the UKB GWAS and the PRS-CS-auto tool with a UK Biobank European ancestry LD reference panel and HapMap3 variants[7]. We calculated individual-level PGSs using the resulting weights in FinnGen study participants with PLINK v2.0[8]. We evaluated the associations of the PAT PGS with cardiovascular diseases registered at any point before or after DNA sampling as outcomes, using logistic regression models with sex, age at the end of study follow-up or death, genomic principal components 1–5 and the genotyping array as basic covariates.

### Supplementary Results

#### Genomic inflation and LD Score Regression Intercept

The genomic control inflation statistic (lambda) of the genome-wide association study of PAT in UKB was modestly elevated at 1.096 (Supplementary Figure 7), but the linkage disequilibrium score regression intercept was 1.00, consistent with true polygenic signal. Similarly, in the meta-analysis of the UKB GWAS and a previously published PAT GWAS meta-analysis[6], the genomic inflation factor was 1.103 and the LD score regression intercept was 1.0105 (SE 0.0075).

### Supplementary Figures

#### Supplementary Figure 1: Study flowchart

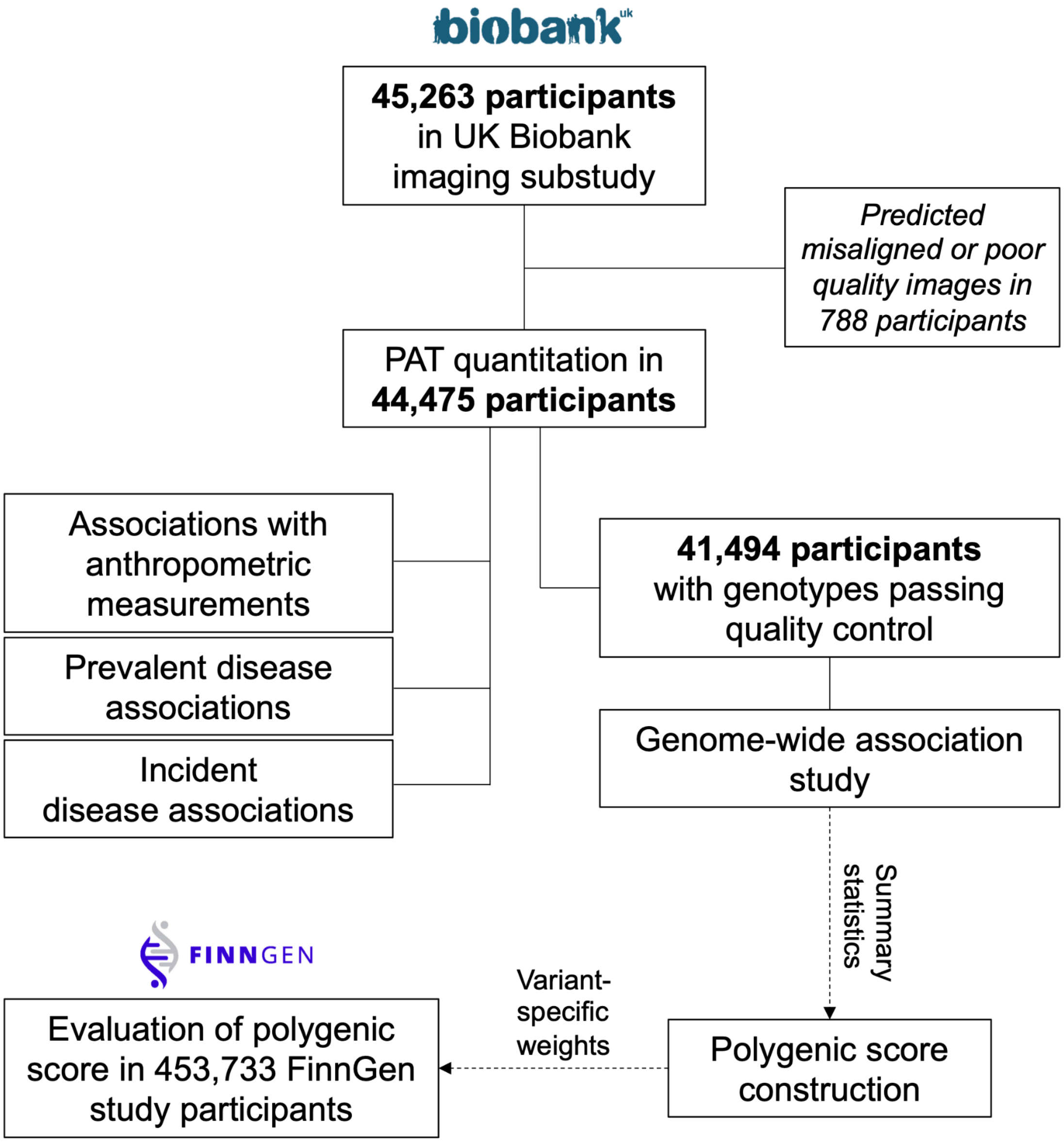

Supplementary Figure 2: The associations of pericardial adipose tissue with age, sex, and anthropometric measurements

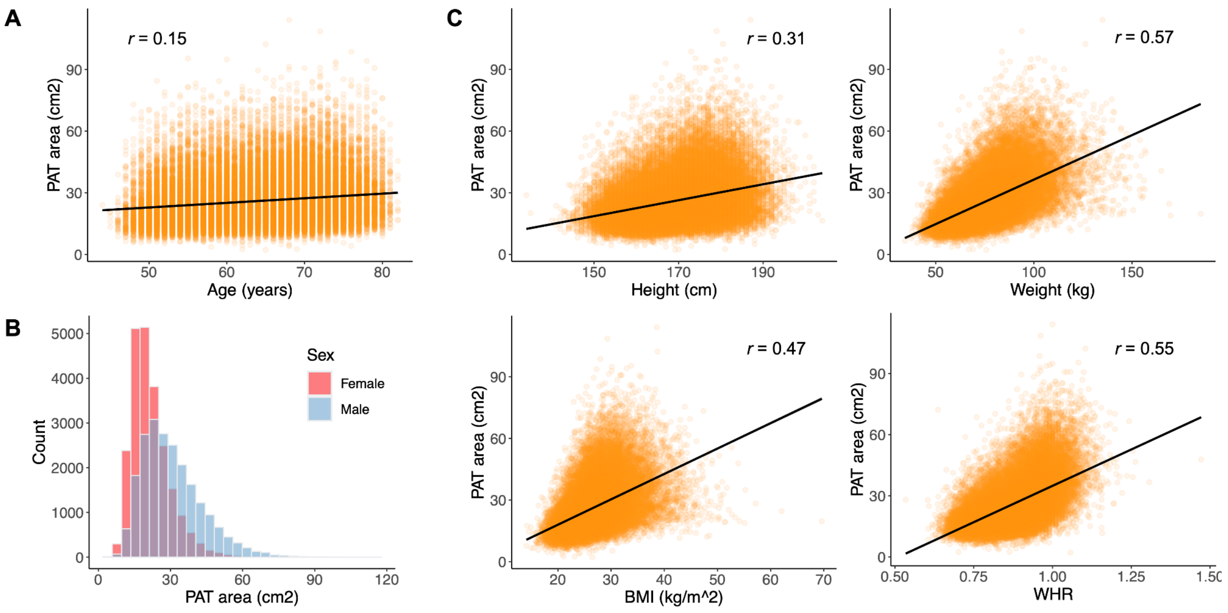


A. The association of PAT with age during the imaging visit. B. The distribution of PAT in women (red) and men (blue). C. The correlations of anthropometric measurements with pericardial adipose tissue. Pearson's correlation coefficient is shown for all pairs of continuous traits. Abbreviations: BMI = body mass index, PAT = pericardial adipose tissue, WHR = waist to hip ratio.

Supplementary Figure 3: The associations of pericardial adipose tissue with incident diseases

#### A. Adjusted for basic covariates

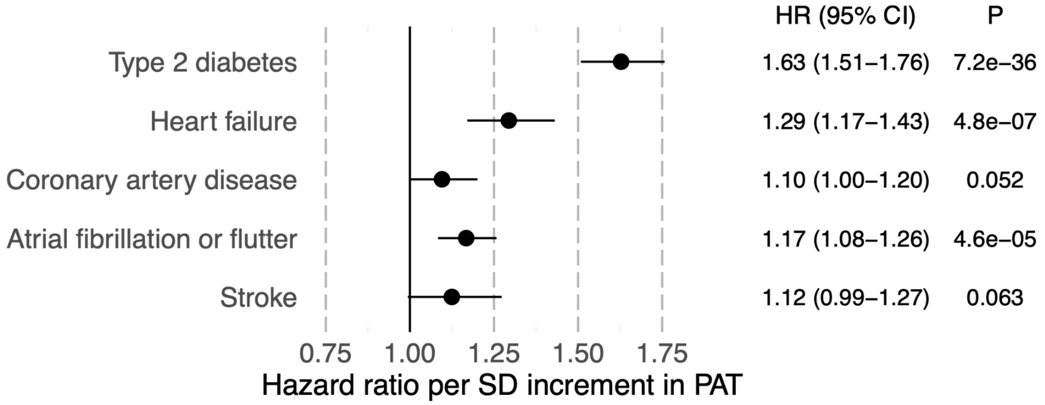


#### B. Adjusted for basic covariates and BMI

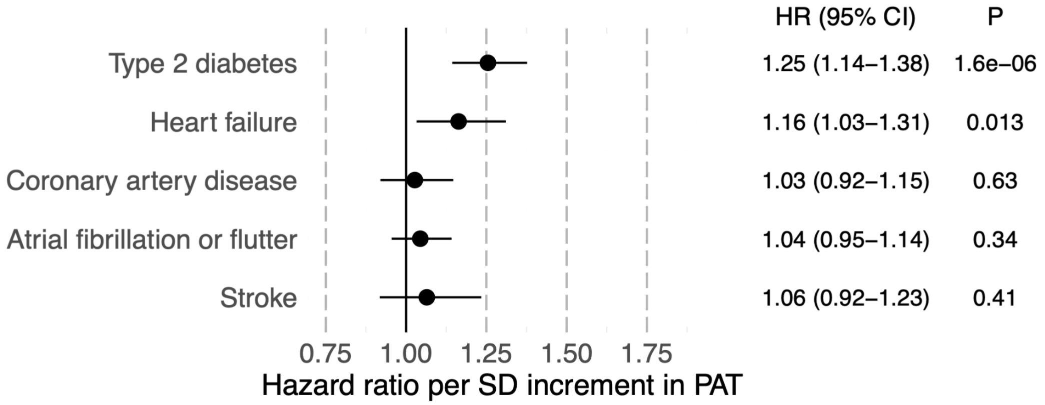


The associations of PAT with incident diseases were tested using Cox proportional hazards models with time from imaging to diagnosis or censoring as the outcome and PAT (standard deviation scaled), sex, and age as the predictors. Body mass index (BMI) was included as an additional covariate in BMI-adjusted models. Participants with the corresponding prevalent disease at the time of imaging were excluded from analyses. Effect estimates are reported for PAT. CI = confidence interval.

Supplementary Figure 4: The associations of percentile-stratified pericardial adipose tissue with incident diseases

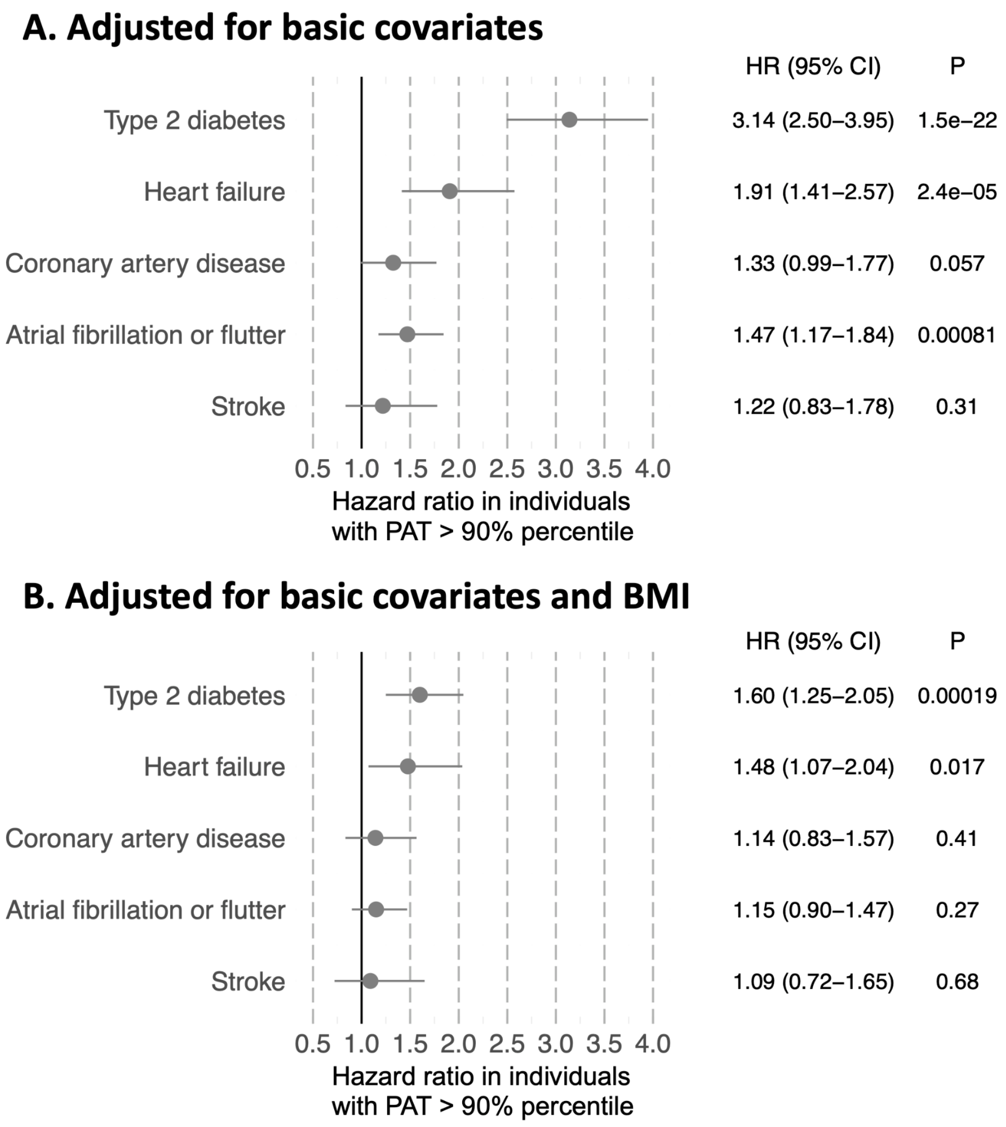


The associations of PAT with incident diseases were tested using Cox proportional hazards models with time from imaging to diagnosis or censoring as the outcome and PAT percentile (over 90th percentile vs. others), sex, and age as the predictors. Participants with the corresponding prevalent disease at the time of imaging were excluded from analyses. Body mass index (BMI) was included as an additional covariate in BMI-adjusted models. Hazard ratios estimates are reported for the PAT strata (> 90th percentile compared with PAT ≤ 90th percentile)

Supplementary Figure 5: Cumulative incidences of cardiovascular diseases stratified by PAT decile at baseline

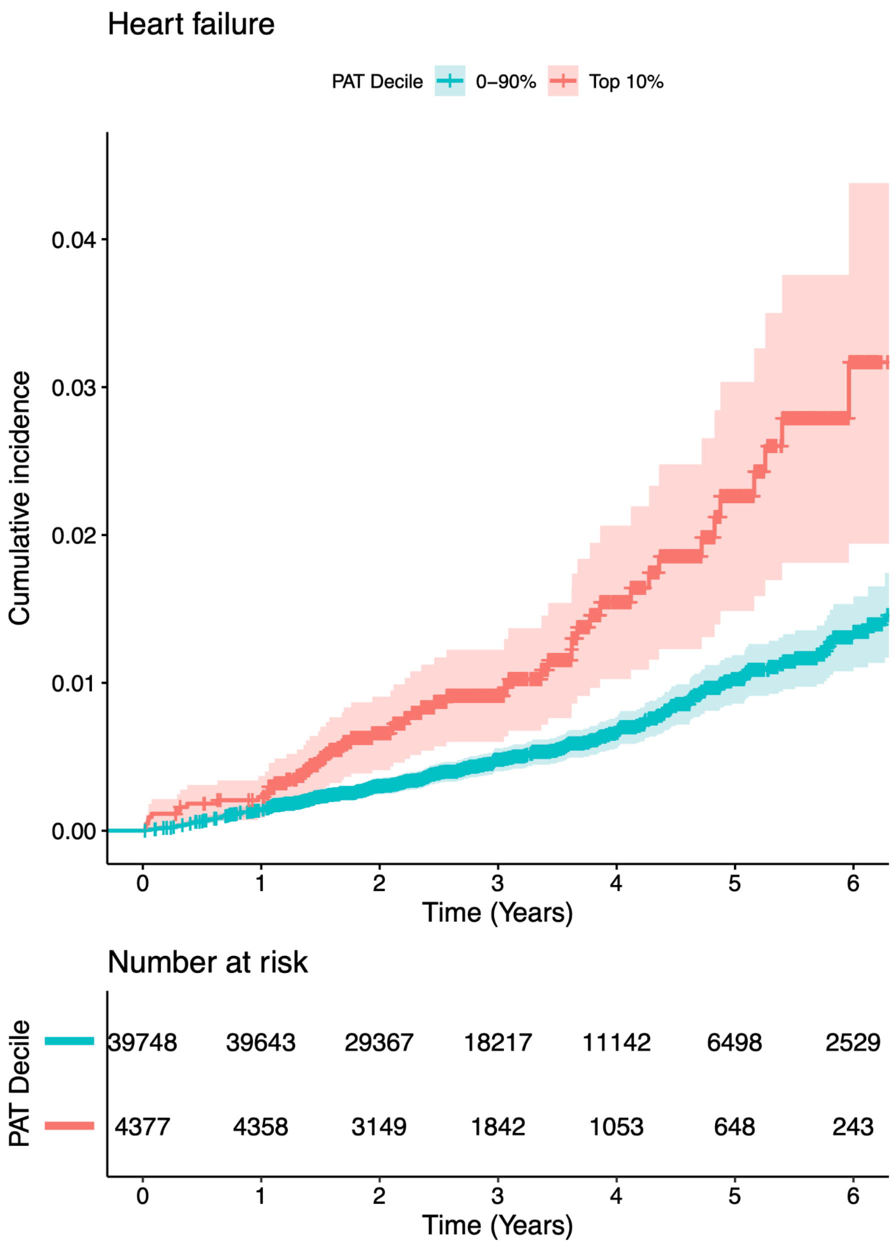

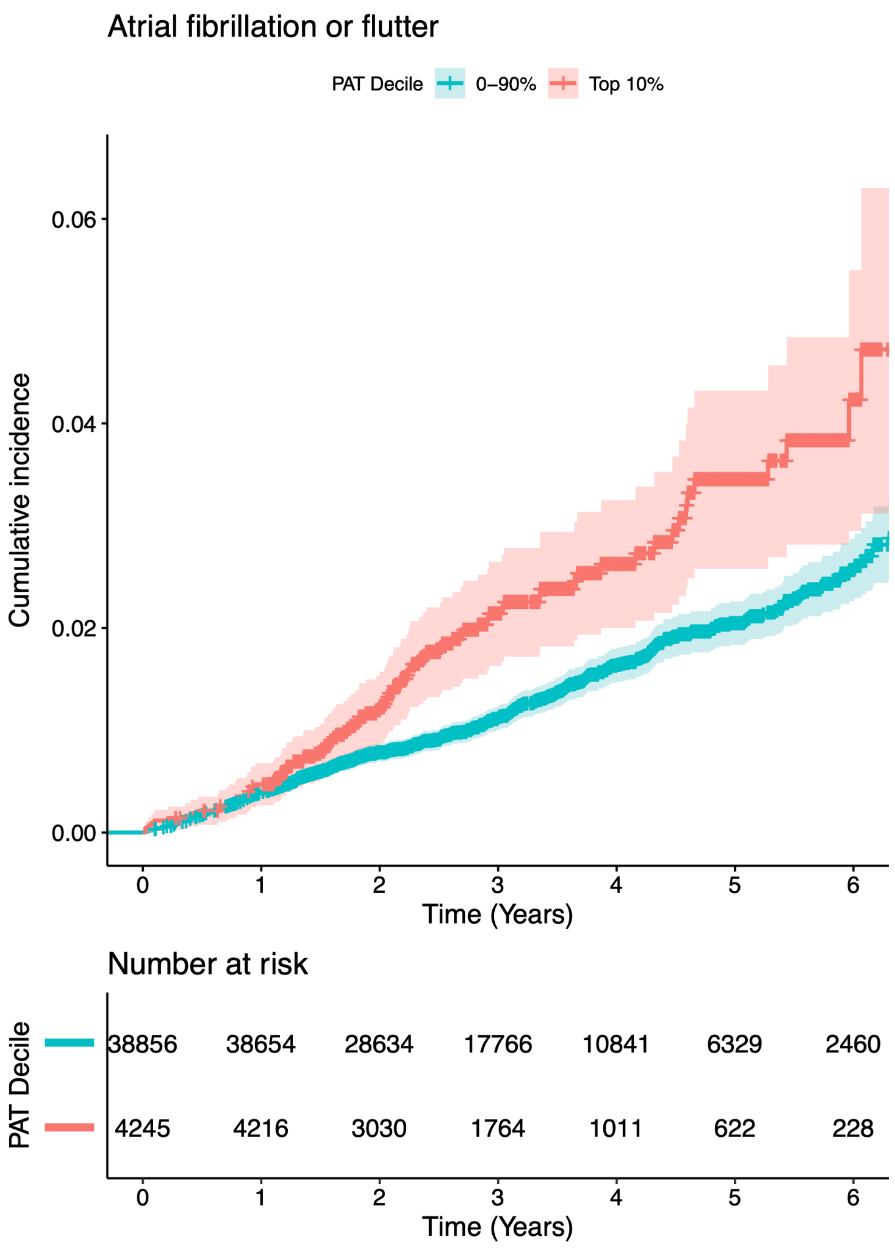
Atrial fibrillation or flutter
PAT Decile
0–90%
Top 10%
Cumulative incidence
0.06
0.04
0.02
0.00
0
1
2
3
4
5
6
Time (Years)
Number at risk
PAT Decile
38856
38654
28634
17766
10841
6329
2460
4245
4216
3030
1764
1011
622
228
0
1
2
3
4
5
6
Time (Years)

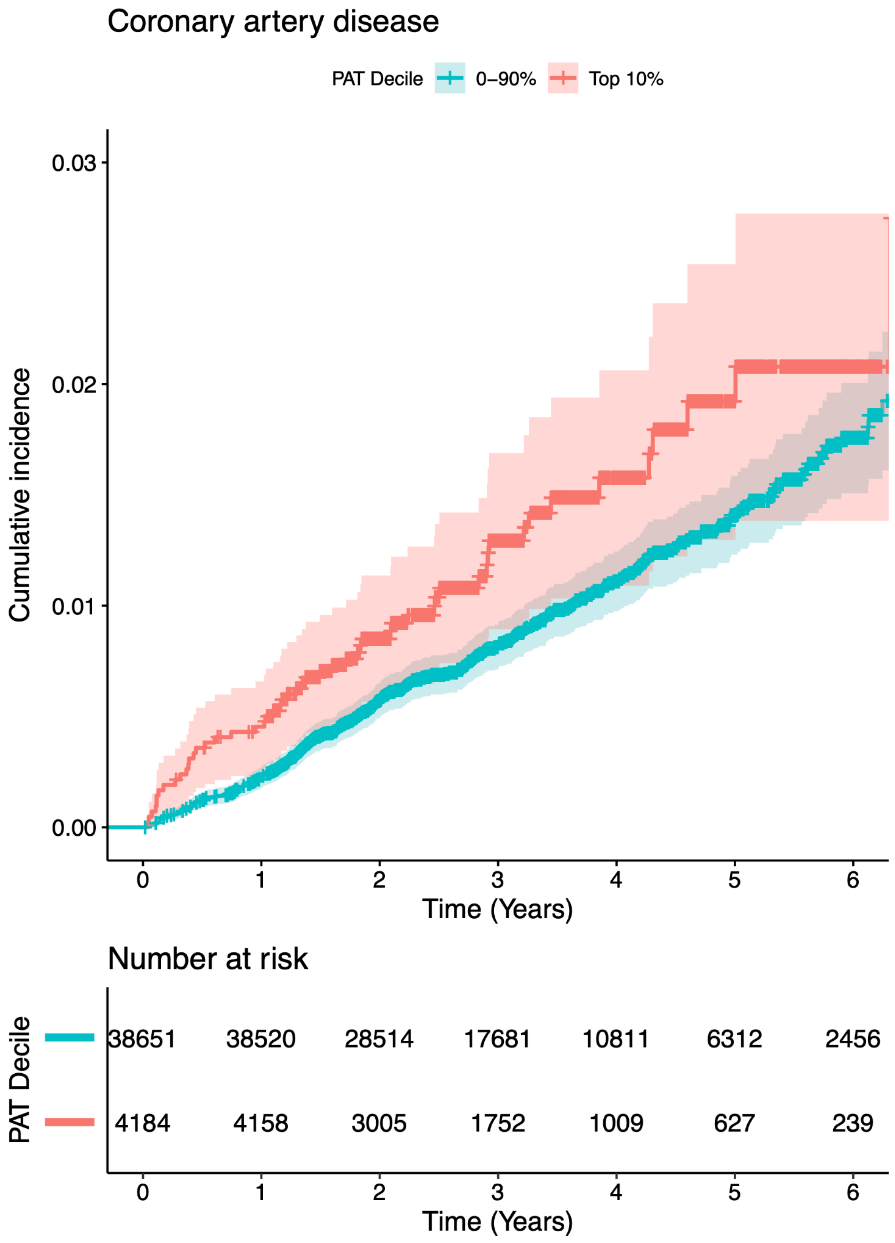
Coronary artery disease
PAT Decile
0–90%
Top 10%
Cumulative incidence
0.03
0.02
0.01
0.00
0
1
2
3
4
5
6
Time (Years)
Number at risk
PAT Decile
38651
38520
28514
17681
10811
6312
2456
4184
4158
3005
1752
1009
627
239
0
1
2
3
4
5
6
Time (Years)

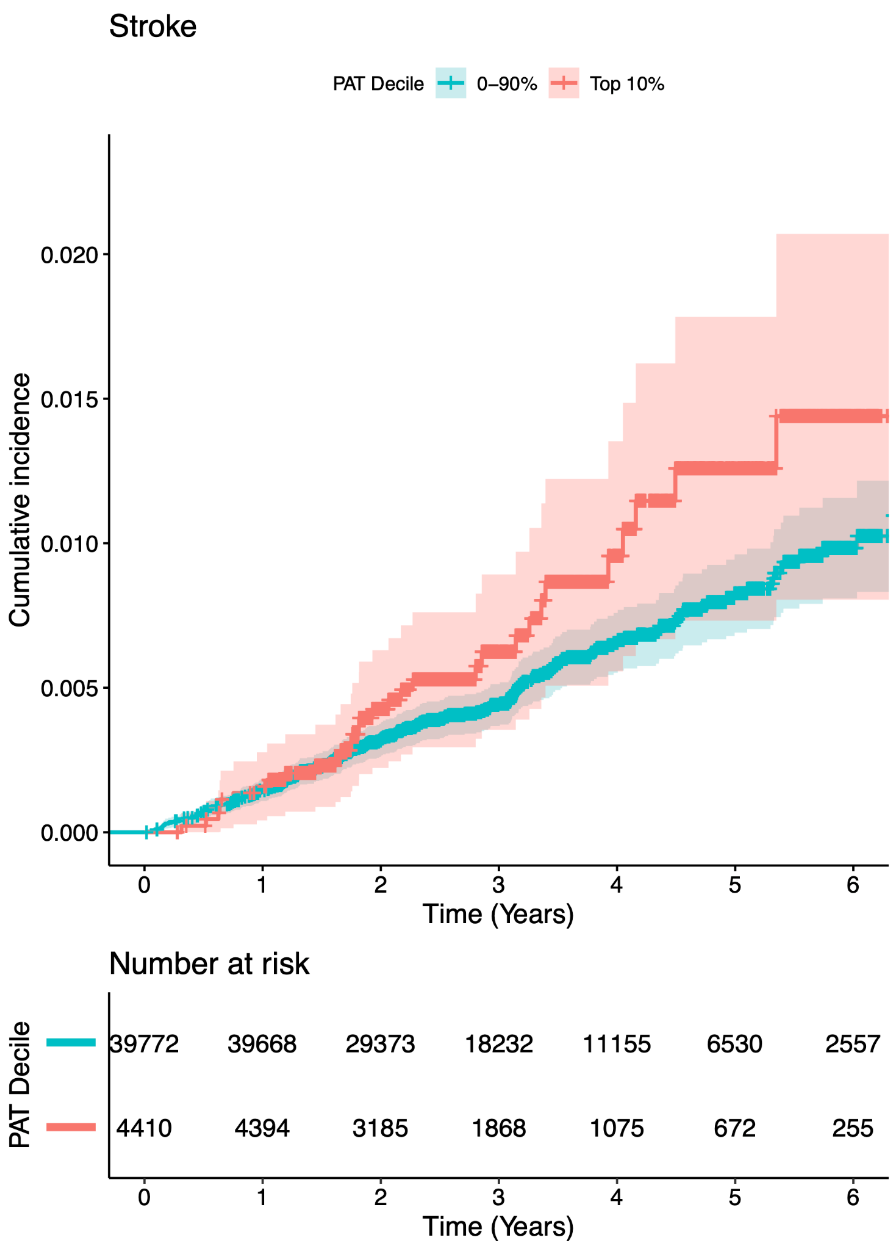
Stroke
PAT Decile
0–90%
Top 10%
Cumulative incidence
0.020
0.015
0.010
0.005
0.000
0
1
2
3
4
5
6
Time (Years)
Number at risk
PAT Decile
39772 39668 29373 18232 11155 6530 2557
4410 4394 3185 1868 1075 672 255
0
1
2
3
4
5
6
Time (Years)

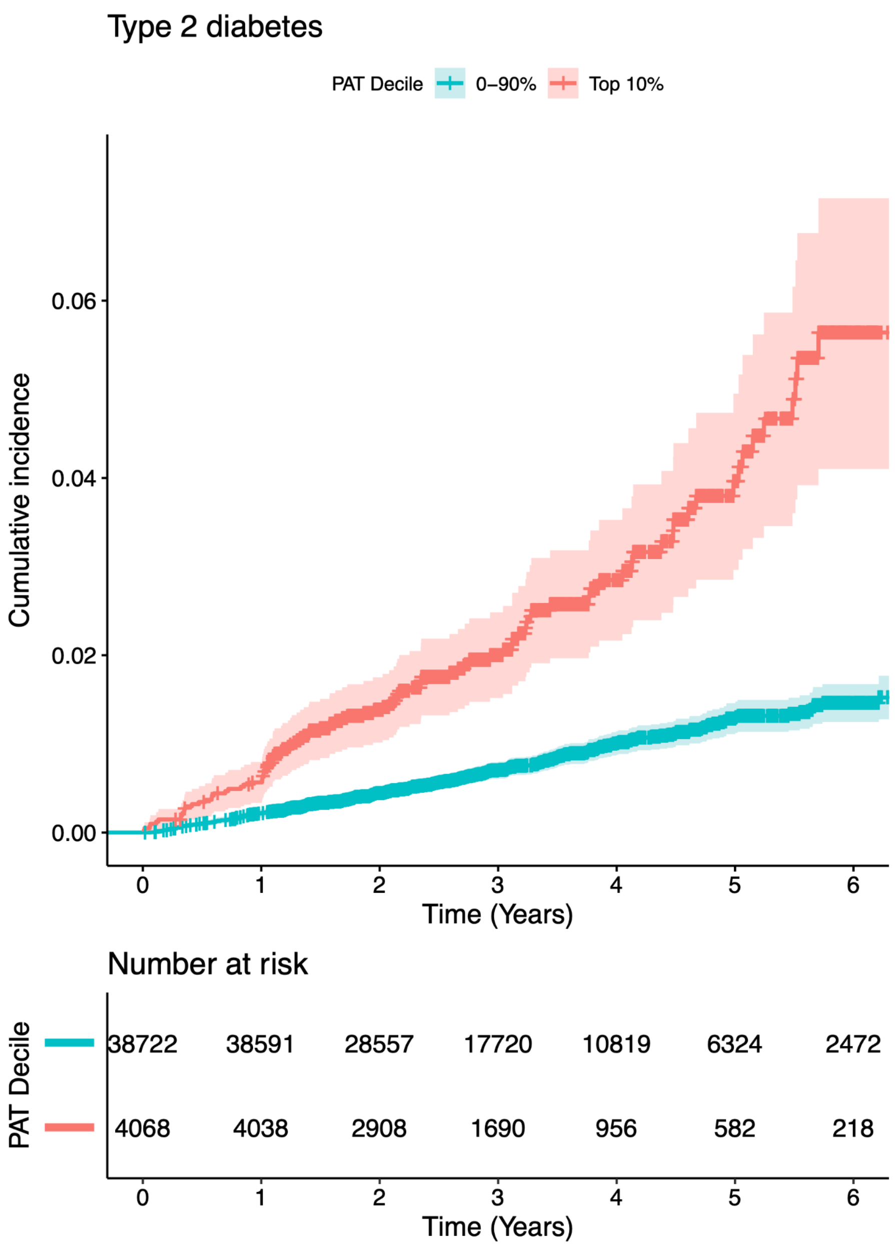
Type 2 diabetes
PAT Decile
0–90%
Top 10%
Cumulative incidence
0.00
0.02
0.04
0.06
0
1
2
3
4
5
6
Time (Years)
Number at risk
PAT Decile
38722 38591 28557 17720 10819 6324 2472
4068 4038 2908 1690 956 582 218
0
1
2
3
4
5
6
Time (Years)

### Supplementary Figure 6: Locus plots from the genome-wide association study of pericardial adipose tissue in UK Biobank

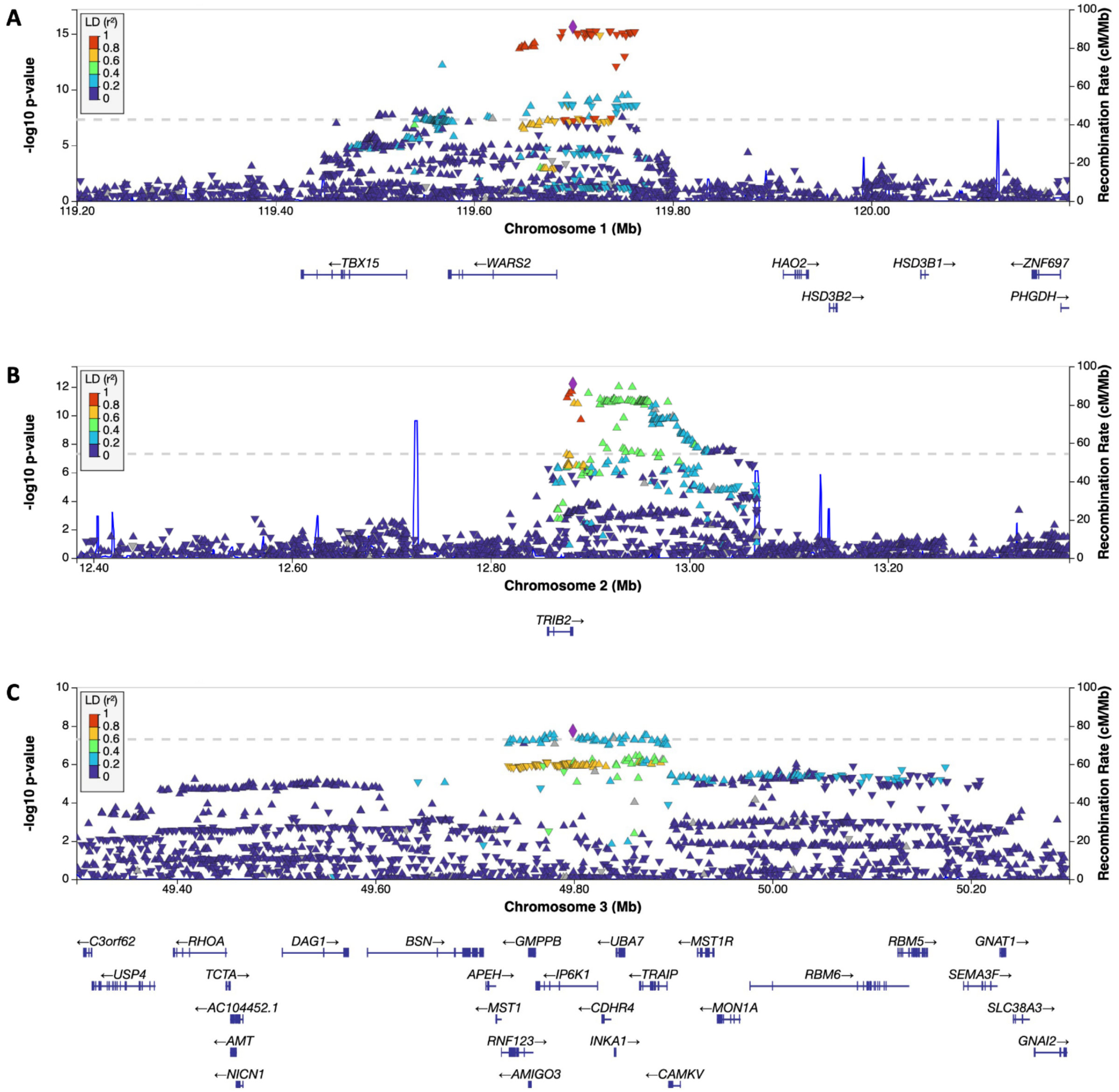

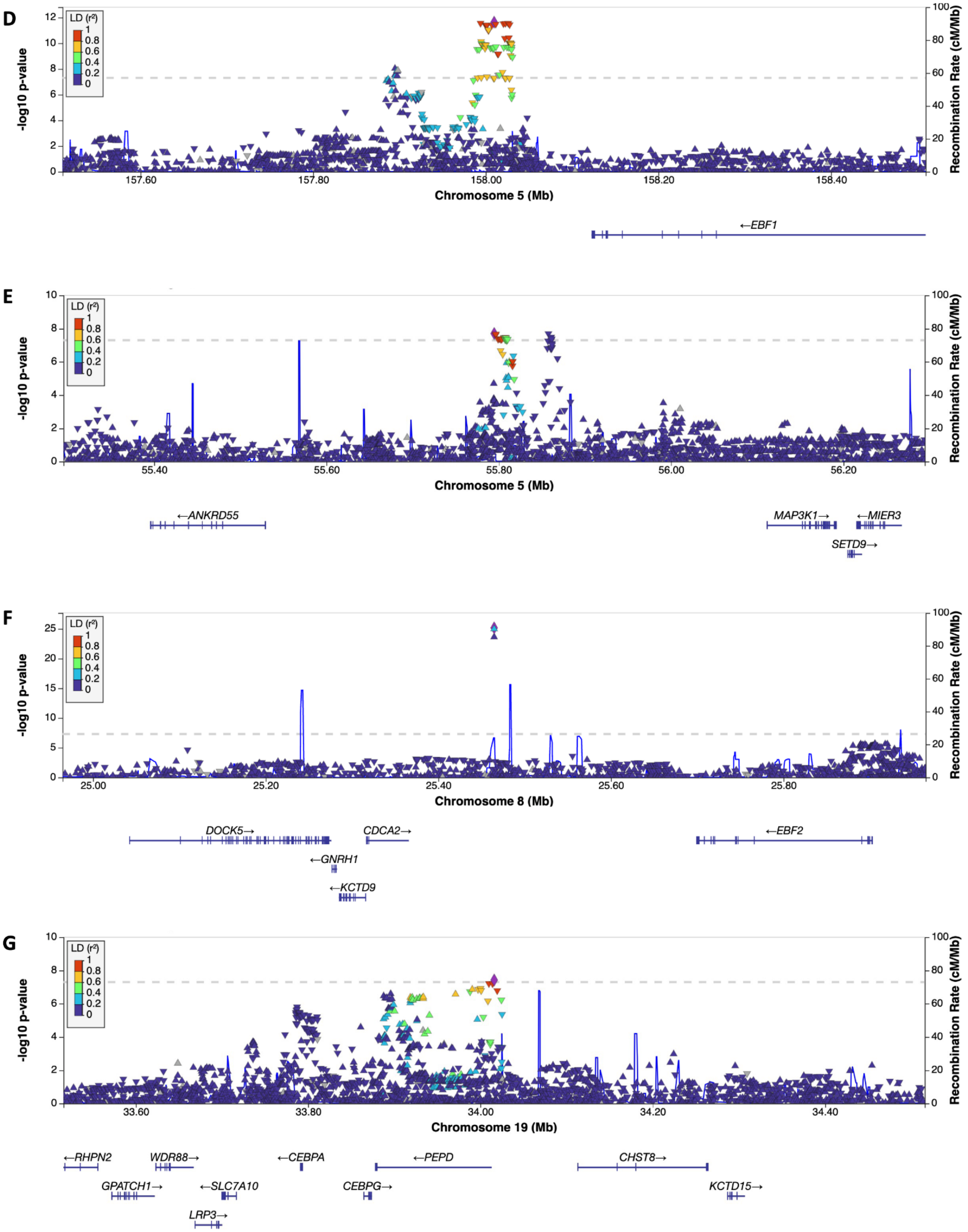

D
-log10 p-value
LD ($r^2$)
1
0.8
0.6
0.4
0.2
0
Recombination Rate (cM/Mb)
157.60
157.80
158.00
158.20
158.40
Chromosome 5 (Mb)
←EBF1
E
-log10 p-value
Recombination Rate (cM/Mb)
55.40
55.60
55.80
56.00
56.20
Chromosome 5 (Mb)
←ANKRD55
MAP3K1→
←MIER3
SETD9→
F
-log10 p-value
Recombination Rate (cM/Mb)
25.00
25.20
25.40
25.60
25.80
Chromosome 8 (Mb)
DOCK5→
CDCA2→
←EBF2
←GNRH1
←KCTD9
G
-log10 p-value
Recombination Rate (cM/Mb)
33.60
33.80
34.00
34.20
34.40
Chromosome 19 (Mb)
←RHPN2
WDR88→
←CEBPA
←PEPD
CHST8→
GPATCH1→
←SLC7A10
CEBPG→
KCTD15→
LRP3→

Supplementary Figure 7: Quantile-quantile plot of p-values from the genome-wide association study of pericardial adipose tissue in UK Biobank

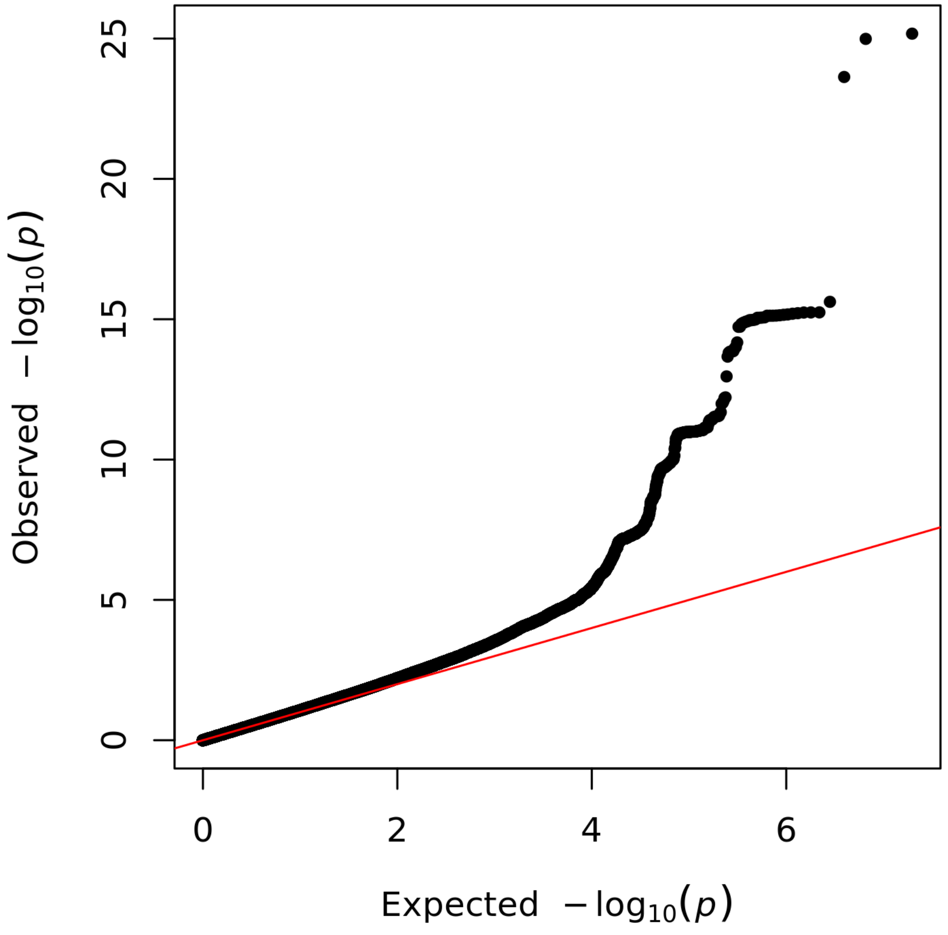


A genome-wide association study of pericardial adipose tissue was performed in 41,494 UK Biobank participants. Expected and observed p-values are shown on the -log10 scale.

#### Supplementary Figure 8: Manhattan plot of the meta-analysis of genome-wide association studies of pericardial adipose tissue

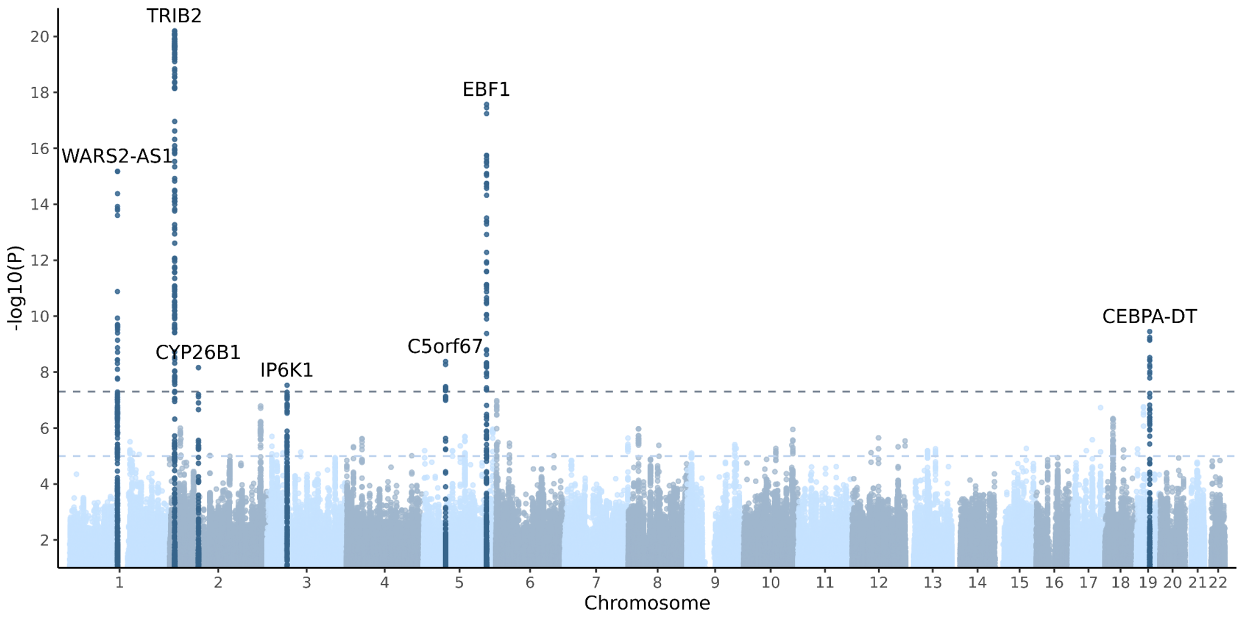


A p-value based sample size weighted meta-analysis of the UK Biobank genome-wide association study and a previously published GWAS meta-analysis by Chu et al (n = 12,204) was performed using METAL, including up to 53,698 total participants. Each variant is plotted as a data point, with the corresponding -$\log_{10}$(P) shown on the y-axis and the genomic position shown on the x-axis grouped by chromosomes. The genome-wide significance threshold (P = $5*10^{-8}$) is shown with a darker dashed line and a suggestive threshold (P = $1*10^{-5}$) is shown with a lighter dashed line. Genomic loci with at least one variant reaching genome-wide significance are labeled with the name of the nearest gene, and all variants within 500 kilobases of the lead variant are coloured in a darker blue for visualization purposes. The y-axis is truncated to only show variants with a P-value ≤ 0.1.

Supplementary Figure 9: Quantile-quantile plot of the meta-analysis of genome-wide association studies of pericardial adipose tissue

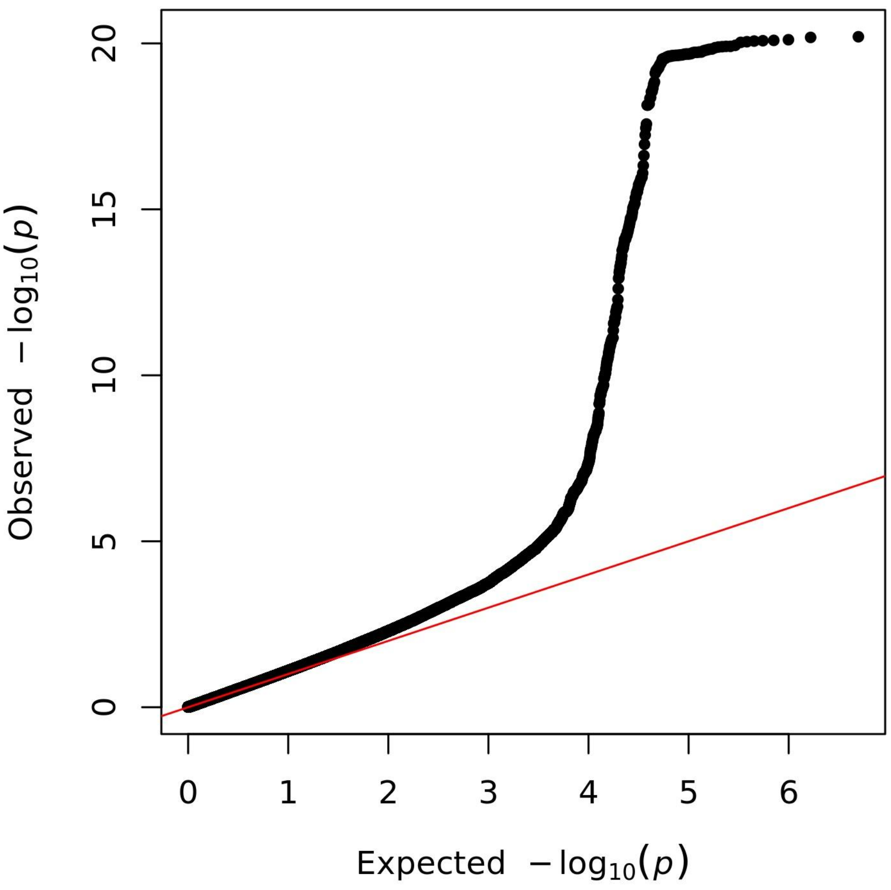


A p-value based sample size weighted meta-analysis of the UK Biobank genome-wide association study and a previously published GWAS meta-analysis by Chu et al (n = 12,204) was performed using METAL, including up to 53,698 total participants. Expected and observed p-values are shown on the -log10 scale.

### Supplementary Tables

#### Supplementary Table 1: Characteristics of the study sample at the time of imaging and stratified by self-reported ethnic background

| | White | Mixed ethnic background | Asian or Asian British | Black or Black British | Chinese | Other ethnic group |
|---|---|---|---|---|---|---|
| N | 43014 | 204 | 482 | 301 | 129 | 226 |
| Male sex (%) | 20768 (48.3) | 73 (35.8) | 307 (63.7) | 138 (45.8) | 49 (38.0) | 101 (44.7) |
| Age (mean (SD)) | 64.21 (7.71) | 59.55 (7.75) | 61.01 (8.44) | 59.15 (7.26) | 59.57 (6.80) | 62.47 (8.04) |
| Height (mean (SD)) | 169.27 (9.24) | 166.41 (8.69) | 166.25 (8.81) | 168.49 (9.51) | 161.60 (7.60) | 164.96 (9.80) |
| Weight (mean (SD)) | 76.15 (15.11) | 75.17 (16.23) | 72.00 (12.59) | 82.07 (16.22) | 62.26 (10.47) | 73.65 (16.09) |
| Body mass index (mean (SD)) | 26.49 (4.37) | 27.09 (5.24) | 25.99 (3.73) | 28.91 (5.30) | 23.79 (3.17) | 26.88 (4.42) |
| Waist circumference (mean (SD)) | 88.37 (12.73) | 85.98 (12.75) | 87.97 (10.76) | 89.74 (12.60) | 78.76 (9.70) | 86.93 (13.30) |
| Hip circumference (mean (SD)) | 100.81 (8.70) | 100.75 (10.64) | 97.62 (7.90) | 103.14 (10.26) | 92.55 (6.66) | 98.85 (9.48) |
| Waist-to-hip ratio (mean (SD)) | 0.88 (0.09) | 0.85 (0.09) | 0.90 (0.08) | 0.87 (0.08) | 0.85 (0.07) | 0.88 (0.09) |
| PAT (mean (SD)) | 26.14 (11.45) | 22.10 (10.02) | 22.95 (9.26) | 20.97 (7.92) | 19.68 (8.05) | 23.12 (10.29) |
| Diabetes type 2 (%) | 1547 (3.6) | 12 (5.9) | 60 (12.4) | 36 (12.0) | 6 (4.7) | 14 (6.2) |
| Heart failure (%) | 304 (0.7) | 2 (1.0) | 5 (1.0) | 1 (0.3) | 0 (0.0) | 3 (1.3) |
| Coronary artery disease (%) | 1573 (3.7) | 4 (2.0) | 36 (7.5) | 6 (2.0) | 1 (0.8) | 11 (4.9) |
| Atrial fibrillation or flutter (%) | 1341 (3.1) | 2 (1.0) | 11 (2.3) | 5 (1.7) | 2 (1.6) | 5 (2.2) |
| Stroke (%) | 454 (1.1) | 1 (0.5) | 5 (1.0) | 3 (1.0) | 1 (0.8) | 1 (0.4) |

Baseline characteristics at the time of imaging are shown for UK Biobank cardiac magnetic resonance imaging substudy participants with automated pericardial adipose tissue (PAT) area quantitation from four-chamber images. Self-reported ancestry was used to stratify participants. Anthropometric measurements were taken during the imaging visit. Prevalent diseases were ascertained based on a combination of self-reported data and International Classification of Diseases codes.

#### Supplementary Table 2: UK Biobank Disease Definitions

| Disease | Field ID | Description | Codes | Included / excluded |
|---|---|---|---|---|
| AF | 20002 | Non-cancer illness code, self-reported | 14,711,483 | Included |
| AF | 20004 | Operation code | 1524 | Included |
| AF | 41202 | Diagnoses - main ICD10 | I48,I48.0,I48.1,I48.2,I48.3,I48.4,I48.9 | Included |
| AF | 41204 | Diagnoses - secondary ICD10 | I48,I48.0,I48.1,I48.2,I48.3,I48.4,I48.9 | Included |
| AF | 40001 | Underlying (primary) cause of death: ICD10 | I48,I48.0,I48.1,I48.2,I48.3,I48.4,I48.9 | Included |
| AF | 40002 | Contributory (secondary) causes of death: ICD10 | I48,I48.0,I48.1,I48.2,I48.3,I48.4,I48.9 | Included |
| AF | 41203 | Diagnoses - main ICD9 | 4273 | Included |
| AF | 41205 | Diagnoses - secondary ICD9 | 4273 | Included |
| AF | 41200 | Operative procedures - main OPCS4 | K57.1,K62.1,K62.2,K62.3,K62.4 | Included |
| AF | 41210 | Operative procedures - secondary OPCS4 | K57.1,K62.1,K62.2,K62.3,K62.4 | Included |
| CAD | 20002 | Non-cancer illness code, self-reported | 1075 | Included |
| CAD | 20004 | Operation code | 107,010,951,523 | Included |
| CAD | 6150 | Vascular/heart problems diagnosed by doctor | 1 | Included |
| CAD | 41202 | Diagnoses - main ICD10 | I21,I21.0,I21.1,I21.2,I21.3,I21.4,I21.9, I22,I22.0,I22.1 | Included |
| CAD | 41202 | Diagnoses - main ICD10 | I22.8,I22.9,I23,I23.0,I23.1,I23.2,I23.3, I23.4,I23.5,I23.6 | Included |
| CAD | 41202 | Diagnoses - main ICD10 | I23.8,I24,I24.0,I24.1,I24.8,I24.9,I25.2 | Included |
| CAD | 41204 | Diagnoses - secondary ICD10 | I21,I21.0,I21.1,I21.2,I21.3,I21.4,I21.9, I22,I22.0,I22.1 | Included |
| CAD | 41204 | Diagnoses - secondary ICD10 | I22.8,I22.9,I23,I23.0,I23.1,I23.2,I23.3, I23.4,I23.5,I23.6 | Included |
| CAD | 41204 | Diagnoses - secondary ICD10 | I23.8,I24,I24.0,I24.1,I24.8,I24.9,I25.2 | Included |
| CAD | 40001 | Underlying (primary) cause of death: ICD10 | I21,I21.0,I21.1,I21.2,I21.3,I21.4,I21.9, I22,I22.0,I22.1 | Included |
| CAD | 40001 | Underlying (primary) cause of death: ICD10 | I22.8,I22.9,I23,I23.0,I23.1,I23.2,I23.3, I23.4,I23.5,I23.6 | Included |

| | | | | |
|---|---|---|---|---|
| CAD | 40001 | Underlying (primary) cause of death: ICD10 | I23.8,I24,I24.0,I24.1,I24.8,I24.9,I25.2 | Included |
| CAD | 40002 | Contributory (secondary) causes of death: ICD10 | I21,I21.0,I21.1,I21.2,I21.3,I21.4,I21.9, I22,I22.0,I22.1 | Included |
| CAD | 40002 | Contributory (secondary) causes of death: ICD10 | I22.8,I22.9,I23,I23.0,I23.1,I23.2,I23.3, I23.4,I23.5,I23.6 | Included |
| CAD | 40002 | Contributory (secondary) causes of death: ICD10 | I23.8,I24,I24.0,I24.1,I24.8,I24.9,I25.2 | Included |
| CAD | 41200 | Operative procedures - main OPCS4 | K40,K40.1,K40.2,K40.3,K40.4,K40.8,K 40.9,K41,K41.1,K41.2 | Included |
| CAD | 41200 | Operative procedures - main OPCS4 | K41.3,K41.4,K41.8,K41.9,K42,K42.1,K 42.2,K42.3,K42.4,K42.8 | Included |
| CAD | 41200 | Operative procedures - main OPCS4 | K42.9,K43,K43.1,K43.2,K43.3,K43.4,K 43.8,K43.9,K44,K44.1 | Included |
| CAD | 41200 | Operative procedures - main OPCS4 | K44.2,K44.8,K44.9,K45.1,K45.2,K45.3, K45.4,K45.5,K45.6,K45.8 | Included |
| CAD | 41200 | Operative procedures - main OPCS4 | K45.9,K46,K46.1,K46.2,K46.3,K46.4,K 46.5,K46.8,K46.9,K49.1 | Included |
| CAD | 41200 | Operative procedures - main OPCS4 | K49.2,K49.3,K49.4,K49.8,K49.9,K50.1, K50.2,K50.4,K75.1,K75.2 | Included |
| CAD | 41200 | Operative procedures - main OPCS4 | K75.3,K75.4,K75.8,K75.9 | Included |
| CAD | 41210 | Operative procedures - secondary OPCS4 | K40,K40.1,K40.2,K40.3,K40.4,K40.8,K 40.9,K41,K41.1,K41.2 | Included |
| CAD | 41210 | Operative procedures - secondary OPCS4 | K41.3,K41.4,K41.8,K41.9,K42,K42.1,K 42.2,K42.3,K42.4,K42.8 | Included |
| CAD | 41210 | Operative procedures - secondary OPCS4 | K42.9,K43,K43.1,K43.2,K43.3,K43.4,K 43.8,K43.9,K44,K44.1 | Included |
| CAD | 41210 | Operative procedures - secondary OPCS4 | K44.2,K44.8,K44.9,K45.1,K45.2,K45.3, K45.4,K45.5,K45.6,K45.8 | Included |
| CAD | 41210 | Operative procedures - secondary OPCS4 | K45.9,K46,K46.1,K46.2,K46.3,K46.4,K 46.5,K46.8,K46.9,K49.1 | Included |
| CAD | 41210 | Operative procedures - secondary OPCS4 | K49.2,K49.3,K49.4,K49.8,K49.9,K50.1, K50.2,K50.4,K75.1,K75.2 | Included |
| CAD | 41210 | Operative procedures - secondary OPCS4 | K75.3,K75.4,K75.8,K75.9 | Included |
| CAD | 41203 | Diagnoses - main ICD9 | 410,410,941,141,194,000,000 | Included |
| CAD | 41205 | Diagnoses - secondary ICD9 | 410,410,941,141,194,000,000 | Included |

| | | | | |
|---|---|---|---|---|
| T2D | 20002 | Non-cancer illness code, self-reported | 1223 | Included |
| T2D | 41202 | Diagnoses - main ICD10 | E11,E11.0,E11.1,E11.2,E11.3,E11.4,E11.5,E11.6,E11.7,E11.8,E11.9 | Included |
| T2D | 41204 | Diagnoses - secondary ICD10 | E11,E11.0,E11.1,E11.2,E11.3,E11.4,E11.5,E11.6,E11.7,E11.8,E11.9 | Included |
| T2D | 40001 | Underlying (primary) cause of death: ICD10 | E11,E11.0,E11.1,E11.2,E11.3,E11.4,E11.5,E11.6,E11.7,E11.8,E11.9 | Included |
| T2D | 40002 | Contributory (secondary) causes of death: ICD10 | E11,E11.0,E11.1,E11.2,E11.3,E11.4,E11.5,E11.6,E11.7,E11.8,E11.9 | Included |
| HF | 20002 | Non-cancer illness code, self-reported | 10,761,079 | Included |
| HF | 41202 | Diagnoses - main ICD10 | I11.0,I13.0,I13.2,I25.5,I42.0,I42.5,I42.8,I42.9,I50,I50.0,I50.1,I50.9 | Included |
| HF | 41204 | Diagnoses - secondary ICD10 | I11.0,I13.0,I13.2,I25.5,I42.0,I42.5,I42.8,I42.9,I50,I50.0,I50.1,I50.9 | Included |
| HF | 40001 | Underlying (primary) cause of death: ICD10 | I11.0,I13.0,I13.2,I25.5,I42.0,I42.5,I42.8,I42.9,I50,I50.0,I50.1,I50.9,4289 | Included |
| HF | 40002 | Contributory (secondary) causes of death: ICD10 | I11.0,I13.0,I13.2,I25.5,I42.0,I42.5,I42.8,I42.9,I50,I50.0,I50.1,I50.9,4289 | Included |
| HF | 41203 | Diagnoses - main ICD9 | 425,442,804,281 | Included |
| HF | 41205 | Diagnoses - secondary ICD9 | 425,442,804,281 | Included |
| HF | 20002 | Non-cancer illness code, self-reported | 1588 | Excluded |
| HF | 41202 | Diagnoses - main ICD10 | I42.1,I42.2 | Excluded |
| HF | 41204 | Diagnoses - secondary ICD10 | I42.1,I42.2 | Excluded |
| HF | 40001 | Underlying (primary) cause of death: ICD10 | I42.1,I42.2 | Excluded |
| HF | 40002 | Contributory (secondary) causes of death: ICD10 | I42.1,I42.2 | Excluded |
| Stroke | 42007 | Source of stroke report | 0,1,2 | Included |

Cardiovascular diseases in UK Biobank were defined using a combination of International Classification of Diseases (ICD) codes, self-report, and procedure codes. AF = atrial fibrillation or flutter, CAD = coronary artery disease, HF = heart failure, T2D = type 2 diabetes.

#### Supplementary Table 3: Prevalent disease association results in the UK Biobank

| Disease | Cases | Controls | Beta | SE | Z | P |
|---|---|---|---|---|---|---|
| Type 2 diabetes | 1679 | 42796 | 0.55795023 | 0.02266754 | 24.6144985 | 6.88E-133 |
| Coronary artery disease | 1634 | 42841 | 0.22275325 | 0.0233208 | 9.55170028 | 1.34E-21 |
| Heart failure | 316 | 44153 | 0.458047 | 0.05158378 | 8.87967091 | 6.95E-19 |
| Atrial fibrillation or flutter | 1368 | 43107 | 0.17963635 | 0.02526582 | 7.10985515 | 1.18E-12 |
| Stroke | 287 | 44188 | 0.09363106 | 0.05410968 | 1.73039391 | 0.08356686 |

In linear regression models, standard deviation scaled pericardial adipose tissue was used as the outcome and prevalent disease status (at the time of imaging), age and sex were used as the predictors. The effect (beta) estimates correspond to the differences in pericardial adipose tissue (in SD units) between individuals with prevalent disease and without prevalent disease.

#### Supplementary Table 4: The associations of pericardial adipose tissue with incident diseases

| Model | Outcome | Cases | Controls | Z | P | HR | HR lower 95% CI | HR upper 95% CI |
|---|---|---|---|---|---|---|---|---|
| Basic | Type 2 diabetes | 394 | 42402 | 12.50 | 7.22E-36 | 1.63 | 1.51 | 1.76 |
| Basic | Heart failure | 281 | 43872 | 5.03 | 4.83E-07 | 1.29 | 1.17 | 1.43 |
| Basic | Atrial fibrillation or flutter | 594 | 42513 | 4.07 | 4.64E-05 | 1.17 | 1.08 | 1.26 |
| Basic | Coronary artery disease | 403 | 42438 | 1.94 | 0.05228927 | 1.10 | 1.00 | 1.20 |
| Basic | Stroke | 238 | 43950 | 1.86 | 0.06274115 | 1.12 | 0.99 | 1.27 |
| BMI-adjusted | Type 2 diabetes | 394 | 42402 | 4.79 | 1.63E-06 | 1.25 | 1.14 | 1.38 |
| BMI-adjusted | Heart failure | 281 | 43872 | 2.49 | 0.01287948 | 1.16 | 1.03 | 1.31 |
| BMI-adjusted | Atrial fibrillation or flutter | 594 | 42513 | 0.95 | 0.34191072 | 1.04 | 0.96 | 1.14 |
| BMI-adjusted | Stroke | 238 | 43950 | 0.83 | 0.40801076 | 1.06 | 0.92 | 1.23 |
| BMI-adjusted | Coronary artery disease | 403 | 42438 | 0.48 | 0.63283322 | 1.03 | 0.92 | 1.15 |

The associations of PAT with incident diseases were tested using Cox proportional hazards models with time from imaging to diagnosis or censoring as the outcome and PAT (standard deviation scaled), sex, and age as the predictors. Body mass index (BMI) was included as an additional covariate in BMI-adjusted models. Participants with the corresponding prevalent disease at the time of imaging were excluded from analyses. Effect estimates are reported for PAT. CI = confidence interval, HR = hazard ratio.

#### Supplementary Table 5: The associations of percentile-stratified pericardial adipose tissue with incident diseases

| Model | Outcome | Cases | Controls | Z | P | HR | HR lower 95% CI | HR upper 95% CI |
|---|---|---|---|---|---|---|---|---|
| Basic | Type 2 diabetes | 394 | 42402 | 9.77 | 1.50E-22 | 3.14 | 2.50 | 3.95 |
| Basic | Heart failure | 281 | 43872 | 4.22 | 2.39E-05 | 1.91 | 1.41 | 2.57 |
| Basic | Atrial fibrillation or flutter | 594 | 42513 | 3.35 | 0.00080946 | 1.47 | 1.17 | 1.84 |
| Basic | Coronary artery disease | 403 | 42438 | 1.91 | 0.05662123 | 1.33 | 0.99 | 1.77 |
| Basic | Stroke | 238 | 43950 | 1.02 | 3.06E-01 | 1.22 | 0.83 | 1.78 |
| BMI-adjusted | Type 2 diabetes | 394 | 42402 | 3.73 | 0.00019376 | 1.60 | 1.25 | 2.05 |
| BMI-adjusted | Heart failure | 281 | 43872 | 2.38 | 0.01741983 | 1.48 | 1.07 | 2.04 |
| BMI-adjusted | Atrial fibrillation or flutter | 594 | 42513 | 1.11 | 0.26531812 | 1.15 | 0.90 | 1.47 |
| BMI-adjusted | Coronary artery disease | 403 | 42438 | 0.83 | 0.40808567 | 1.14 | 0.83 | 1.57 |
| BMI-adjusted | Stroke | 238 | 43950 | 0.41 | 0.68065094 | 1.09 | 0.72 | 1.65 |

The associations of PAT with incident diseases were tested using Cox proportional hazards models with time from imaging to diagnosis or censoring as the outcome and PAT percentile (over 90th percentile vs. others), sex, and age as the predictors. Participants with the corresponding prevalent disease at the time of imaging were excluded from analyses. Body mass index (BMI) was included as an additional covariate in BMI-adjusted models. Effect estimates are reported for the PAT strata (> 90th percentile compared with PAT ≤ 90th percentile). CI = confidence interval, HR = hazard ratio.

Supplementary Table 6: Polygenic priority scores for genes in genome-wide significant loci in the genome-wide association study of pericardial adipose tissue in UK Biobank

| Locus | Min pos | Max pos | ENSGID | Name | PoPS | Start | End | TSS |
|---|---|---|---|---|---|---|---|---|
| 1 | 119199426 | 120199426 | ENSG00000116874 | WARS2 | 1.03888821 | 119573839 | 119683294 | 119683294 |
| 1 | 119199426 | 120199426 | ENSG00000092607 | TBX15 | 0.97657313 | 119425669 | 119532179 | 119532179 |
| 1 | 119199426 | 120199426 | ENSG00000203857 | HSD3B1 | 0.23222331 | 120049821 | 120057681 | 120049821 |
| 1 | 119199426 | 120199426 | ENSG00000143067 | ZNF697 | 0.05501743 | 120162045 | 120190396 | 120190396 |
| 1 | 119199426 | 120199426 | ENSG00000203859 | HSD3B2 | -0.0329964 | 119957554 | 119965658 | 119957554 |
| 1 | 119199426 | 120199426 | ENSG00000116882 | HAO2 | -0.2148152 | 119911402 | 119936753 | 119911402 |
| 2 | 12382822 | 13382822 | ENSG00000071575 | TRIB2 | 1.58198676 | 12857015 | 12882860 | 12857015 |
| 3 | 49299046 | 50299046 | ENSG00000004534 | RBM6 | 0.60043145 | 49977440 | 50137478 | 49977440 |
| 3 | 49299046 | 50299046 | ENSG00000233276 | GPX1 | 0.52725384 | 49394609 | 49396033 | 49396033 |
| 3 | 49299046 | 50299046 | ENSG00000173402 | DAG1 | 0.49902449 | 49506146 | 49573048 | 49506146 |
| 3 | 49299046 | 50299046 | ENSG00000001617 | SEMA3F | 0.49041147 | 50192478 | 50226508 | 50192478 |
| 3 | 49299046 | 50299046 | ENSG00000067560 | RHOA | 0.4330987 | 49396578 | 49450431 | 49450431 |
| 3 | 49299046 | 50299046 | ENSG00000164068 | RNF123 | 0.42887217 | 49726932 | 49758962 | 49726932 |
| 3 | 49299046 | 50299046 | ENSG00000176095 | IP6K1 | 0.36463148 | 49761727 | 49823975 | 49823975 |
| 3 | 49299046 | 50299046 | ENSG00000003756 | RBM5 | 0.32854133 | 50126341 | 50156454 | 50126341 |
| 3 | 49299046 | 50299046 | ENSG00000145020 | AMT | 0.22330205 | 49454211 | 49460186 | 49460186 |
| 3 | 49299046 | 50299046 | ENSG00000188315 | C3orf62 | 0.21420054 | 49306035 | 49315342 | 49315342 |
| 3 | 49299046 | 50299046 | ENSG00000164078 | MST1R | 0.17829801 | 49924435 | 49941299 | 49941299 |
| 3 | 49299046 | 50299046 | ENSG00000145022 | TCTA | 0.17811643 | 49449639 | 49453908 | 49449639 |

| | | | | | | | | |
|---|---|---|---|---|---|---|---|---|
| 3 | 49299046 | 50299046 | ENSG00000164077 | MON1A | 0.17742103 | 49946302 | 49967606 | 49967606 |
| 3 | 49299046 | 50299046 | ENSG00000164062 | APEH | 0.16471345 | 49711435 | 49721396 | 49711435 |
| 3 | 49299046 | 50299046 | ENSG00000164076 | CAMKV | 0.12723149 | 49895421 | 49907655 | 49907655 |
| 3 | 49299046 | 50299046 | ENSG00000182179 | UBA7 | 0.12413841 | 49842640 | 49851379 | 49851379 |
| 3 | 49299046 | 50299046 | ENSG00000114316 | USP4 | 0.11628078 | 49315264 | 49378145 | 49378145 |
| 3 | 49299046 | 50299046 | ENSG00000173531 | MST1 | 0.08499937 | 49721380 | 49726934 | 49726934 |
| 3 | 49299046 | 50299046 | ENSG00000164061 | BSN | 0.0497476 | 49591922 | 49708978 | 49591922 |
| 3 | 49299046 | 50299046 | ENSG00000187492 | CDHR4 | 0.04815737 | 49828165 | 49837268 | 49837268 |
| 3 | 49299046 | 50299046 | ENSG00000185614 | FAM212A | 0.0455302 | 49840687 | 49842463 | 49840687 |
| 3 | 49299046 | 50299046 | ENSG00000173540 | GMPPB | 0.03592265 | 49754277 | 49761384 | 49761384 |
| 3 | 49299046 | 50299046 | ENSG00000176020 | AMIGO3 | -0.0764481 | 49754267 | 49761349 | 49761349 |
| 3 | 49299046 | 50299046 | ENSG00000114353 | GNAI2 | -0.0862229 | 50263724 | 50296787 | 50263724 |
| 3 | 49299046 | 50299046 | ENSG00000145029 | NICN1 | -0.1067876 | 49460379 | 49466759 | 49466759 |
| 3 | 49299046 | 50299046 | ENSG00000183763 | TRAIP | -0.1813272 | 49866034 | 49894007 | 49894007 |
| 3 | 49299046 | 50299046 | ENSG00000114349 | GNAT1 | -0.3860586 | 50229045 | 50233949 | 50229045 |
| 4 | 157509651 | 158509651 | ENSG00000164330 | EBF1 | 0.61142758 | 158122928 | 158526769 | 158526769 |
| 5 | 55294632 | 56294632 | ENSG00000155545 | MIER3 | 0.47698095 | 56215429 | 56267502 | 56267502 |
| 5 | 55294632 | 56294632 | ENSG00000155542 | SETD9 | 0.07370149 | 56205087 | 56221359 | 56205087 |
| 5 | 55294632 | 56294632 | ENSG00000095015 | MAP3K1 | -0.1125205 | 56111401 | 56191979 | 56111401 |
| 5 | 55294632 | 56294632 | ENSG00000164512 | ANKRD55 | -0.2717545 | 55395507 | 55529186 | 55529186 |
| 6 | 24964670 | 25964670 | ENSG00000221818 | EBF2 | 1.86841812 | 25699246 | 25902913 | 25902913 |
| 6 | 24964670 | 25964670 | ENSG00000147459 | DOCK5 | 1.01477449 | 25042238 | 25275598 | 25042238 |
| 6 | 24964670 | 25964670 | ENSG00000104756 | KCTD9 | 0.36411649 | 25285366 | 25315992 | 25315992 |
| 6 | 24964670 | 25964670 | ENSG00000147437 | GNRH1 | 0.24394595 | 25276776 | 25282170 | 25282170 |

| | | | | | | | | |
|---|---|---|---|---|---|---|---|---|
| 6 | 24964670 | 25964670 | ENSG00000184661 | CDCA2 | -0.2873648 | 25316513 | 25365436 | 25316513 |
| 7 | 33515904 | 34515904 | ENSG00000245848 | CEBPA | 2.19365014 | 33790840 | 33793470 | 33793470 |
| 7 | 33515904 | 34515904 | ENSG00000124299 | PEPD | 0.73815334 | 33877856 | 34012700 | 34012700 |
| 7 | 33515904 | 34515904 | ENSG00000153885 | KCTD15 | 0.37410233 | 34286838 | 34306668 | 34286838 |
| 7 | 33515904 | 34515904 | ENSG00000153879 | CEBPG | 0.2954409 | 33864236 | 33873592 | 33864236 |
| 7 | 33515904 | 34515904 | ENSG00000076650 | GPATCH1 | 0.17831111 | 33571786 | 33621448 | 33571786 |
| 7 | 33515904 | 34515904 | ENSG00000131941 | RHPN2 | 0.14773468 | 33469499 | 33555794 | 33555794 |
| 7 | 33515904 | 34515904 | ENSG00000130881 | LRP3 | 0.03669161 | 33668509 | 33698694 | 33668509 |
| 7 | 33515904 | 34515904 | ENSG00000166359 | WDR88 | -0.0535222 | 33622996 | 33666701 | 33622996 |
| 7 | 33515904 | 34515904 | ENSG00000130876 | SLC7A10 | -0.1138438 | 33699570 | 33716756 | 33716756 |
| 7 | 33515904 | 34515904 | ENSG00000124302 | CHST8 | -0.2077671 | 34112861 | 34264414 | 34112861 |

Polygenic Priority Scores (PoPS) were calculated for all genes within +/- 500,000kb of the lead variant in each genome-wide significant locus. “Min pos” and “Max pos” correspond to the locus boundaries. Start and End correspond to the regions of each gene. TSS = Transcription Start Site.

#### Supplementary Table 7: Genetic correlations between pericardial adipose tissue and anthropometric traits

| Phenotype 1 | Phenotype 2 | rg | se | z | p |
|---|---|---|---|---|---|
| PAT | Height | 0.05 | 0.05 | 1.00 | 0.32 |
| PAT | Weight | 0.48 | 0.04 | 11.37 | 5.93E-30 |
| PAT | BMI | 0.50 | 0.04 | 11.40 | 4.29E-30 |
| PAT | WHR | 0.60 | 0.05 | 11.44 | 2.69E-30 |

Genetic correlations between pericardial adipose tissue (PAT) and anthropometric measurements were calculated based on the summary statistics from genome-wide association studies in the UKB imaging substudy, using LD Score Regression with HapMap3 variants and a European ancestry reference panel. rg = genetic correlation, se = standard error.

#### Supplementary Table 8: Lead variants from the meta-analysis of genome-wide association studies of pericardial adipose tissue

| CHROM | POS | ID | EA | NEA | Nearest gene | Function | P | N | EA AF (UKB) |
|---|---|---|---|---|---|---|---|---|---|
| 1 | 119736529 | rs10802081 | C | T | WARS2-AS1 | ncRNA_intronic | 6.61E-16 | 53081 | 0.674132 |
| 2 | 12951752 | rs2113814 | T | C | TRIB2 | intergenic | 6.31E-21 | 53080 | 0.495936 |
| 2 | 72178287 | rs908611 | T | C | CYP26B1 | intergenic | 6.92E-09 | 52569 | 0.600742 |
| 3 | 49823024 | rs11920900 | C | G | IP6K1 | intronic | 2.95E-08 | 48949 | 0.316931 |
| 5 | 55860781 | rs3936511 | G | A | C5orf67 | ncRNA_intronic | 4.17E-09 | 53096 | 0.190317 |
| 5 | 158025983 | rs748510 | C | T | EBF1 | intergenic | 2.69E-18 | 53092 | 0.237497 |
| 19 | 33795241 | rs16967952 | A | G | CEBPA-DT | ncRNA_exonic | 3.55E-10 | 51805 | 0.134691 |

A p-value based sample size weighted meta-analysis of the UK Biobank genome-wide association study and a previously published GWAS meta-analysis by Chu et al (n = 12,204) was performed using METAL, including up to 53,698 total participants. Lead variants, nearest genes and variant functions are shown. Variant allele frequencies (AF) are based on UK Biobank data only. CHROM = chromosome, POS = position in the GRCh37 build.

#### Supplementary Table 9: FinnGen participant characteristics

| **Characteristic** | **Overall** | **Females** | **Males** |
|---|---|---|---|
| N | 453733 | 254618 | 199115 |
| Age at genotyping (mean (SD)) | 52.92 (17.96) | 51.25 (18.15) | 55.07 (17.47) |
| Age at death or end of follow-up (mean (SD)) | 60.51 (17.97) | 58.05 (18.21) | 63.67 (17.15) |
| BMI (mean (SD)) | 27.31 (5.54) | 27.27 (6.04) | 27.34 (4.93) |
| Male sex (%) | 199115 (43.9) | 0 (0.0) | 199115 (100.0) |
| Type 2 diabetes (%) | 79190 (17.7) | 35381 (14.1) | 43809 (22.3) |
| Atrial fibrillation or flutter (%) | 55853 (19.4) | 21088 (12.5) | 34765 (29.2) |
| Heart failure (%) | 33250 (7.3) | 12476 (4.9) | 20774 (10.4) |
| Coronary heart disease (%) | 51098 (11.3) | 15028 (5.9) | 36070 (18.1) |
| Stroke (%) | 30521 (6.9) | 11923 (4.8) | 18598 (9.7) |

Sample characteristics are shown for 453,733 individuals from data freeze 11. Cardiovascular diseases were defined using a combination of International Classification of Diseases (ICD) codes from specialist inpatient, outpatient and cause-of-death registries, procedure codes, and medication reimbursement codes. Sex, age at genotyping, and body mass index (BMI) were ascertained at study recruitment. Disease-specific outcomes include both prevalent and incident diseases before the end of the follow-up.

#### Supplementary Table 10: The predictive utility of a polygenic score for pericardial adipose tissue in FinnGen

| **Model** | **Outcome** | **Beta** | **SE** | **OR** | **OR lower 95% CI** | **OR upper 95% CI** | **z** | **P** | **Cases** | **Controls** |
|---|---|---|---|---|---|---|---|---|---|---|
| Basic | Stroke | 0.02 | 0.01 | 1.02 | 1.01 | 1.03 | 2.92 | 0.0035 | 30521 | 408788 |
| Basic | Heart failure | 0.05 | 0.01 | 1.05 | 1.04 | 1.06 | 7.83 | 4.79E-15 | 33250 | 420483 |
| Basic | Type 2 diabetes | 0.06 | 0.00 | 1.06 | 1.05 | 1.07 | 13.94 | 3.57E-44 | 79190 | 369038 |
| Basic | Atrial fibrillation or flutter | 0.04 | 0.01 | 1.04 | 1.03 | 1.06 | 6.85 | 7.60E-12 | 55853 | 231952 |
| Basic | Coronary artery disease | 0.03 | 0.01 | 1.03 | 1.02 | 1.04 | 6.42 | 1.37E-10 | 51098 | 402635 |
| BMI-adjusted | Stroke | 0.02 | 0.01 | 1.02 | 1.01 | 1.03 | 2.81 | 0.0050 | 30521 | 408788 |
| BMI-adjusted | Heart failure | 0.03 | 0.01 | 1.03 | 1.02 | 1.05 | 4.88 | 1.06E-06 | 33250 | 420483 |
| BMI-adjusted | Type 2 diabetes | 0.02 | 0.01 | 1.02 | 1.01 | 1.03 | 4.49 | 6.98E-06 | 79190 | 369038 |
| BMI-adjusted | Atrial fibrillation or flutter | 0.02 | 0.01 | 1.02 | 1.01 | 1.04 | 2.75 | 0.0060 | 55853 | 231952 |
| BMI-adjusted | Coronary artery disease | 0.03 | 0.01 | 1.03 | 1.02 | 1.04 | 4.87 | 1.10E-06 | 51098 | 402635 |

A polygenic score (PGS) for pericardial adipose tissue (PAT) was constructed using PRS-CS based on summary statistics from the genome-wide association study of PAT in UK Biobank and subsequently applied to 453,733 participants in the FinnGen study. The associations of the PAT PGS with cardiovascular diseases were evaluated using logistic regression models with sex, age at the end of study follow-up or death, genomic principal components 1–5 and the genotyping array as basic covariates. In body mass index (BMI) adjusted models, BMI was included as an additional covariate. Effect estimates are shown per SD increment in PAT PGS. CI = confidence interval, OR = odds ratio.
